# Radiomics of the Airway (RadAr): Multi-Scale Airway Phenotyping for Disease Characterization on Routine CT Imaging

**DOI:** 10.64898/2026.07.19.26358441

**Authors:** Pushkar Mutha, Juyoung Lee, George Lucas Silva, Bastiaan Dreihuys, Zachary Healy, David Mummy, Bhavika Kaul, Sundaresh Ram, Rabindra Tirouvanziam, Lokesh Guglani, Anant Madabhushi

**Author notes:** **Corresponding Author:** Anant Madabhushi, PhD Robert W Woodruff Professor, The Wallace H. Coulter Department of Biomedical Engineering Georgia Institute of Technology and Emory University.

## Abstract

**Background:** Airway remodeling is a convergent feature across respiratory diseases, yet current quantitative CT tools quantify a small number of prespecified structural abnormalities of the airway tree. Radiomics of the Airway (RadAr) derives interpretable, multi-scale airway measurements from chest CT to characterize airway deformation and discover quantitative imaging biomarkers from routine chest CT.

**Methods:** RadAr extracts over 400 multi-scale, interpretable airway measurements across lobes or generations capturing luminal dimensions, tapering, architectural distortion, and global morphology. It was evaluated in N=1331 patients across four settings: 63-week mortality prediction in fibrotic interstitial lung disease (fILD), COVID-19 severity prediction, structure-function association in progressive pulmonary fibrosis (PPF) and structure-inflammation markers in pediatric cystic fibrosis (CF). Unsupervised clustering was used to identify airway phenotypes across the fILD and COVID-19 cohorts.

**Results:** In fILD, lower-lobe architectural distortion was associated with mortality (balanced accuracy 0·654). In COVID-19, severe disease was independently associated with luminal dilation (AUC 0·719, odds ratio 2·32, p=0·017). In PPF, airway phenotypes correlated with forced vital capacity (ρ=0·83), mid-expiratory flow (ρ=0·87), and ¹²⁹Xe MRI alveolar gas exchange impairment (ρ=0·70). In pediatric CF, reduced tapering and increased cylindricity were associated with prior exacerbations and bronchoalveolar lavage neutrophilia (ρ=-0·64 to -0·78). Five phenotypes were identified from extensive, tapered airway trees to sparse, dilated, thick-walled, tortuous trees, with increasing COVID-19 severity and fILD mortality across this spectrum.

**Conclusion:** RadAr identified interpretable, disease-specific airway phenotypes associated with function and outcomes across restrictive, obstructive, and mixed lung diseases in adult and pediatric settings. These findings establish a framework for discovery and development of quantitative airway biomarkers for patient stratification, disease monitoring, or imaging endpoint development in pulmonary trials.

## Introduction

Across various respiratory diseases, inflammation, infection, mucus impaction, and fibrosis cause significant remodeling of the airway wall, lumen, and surrounding parenchyma. In fibrotic interstitial lung diseases (fILD), traction bronchiectasis and architectural distortion on computed tomography (CT) are associated with disease progression and survival^1^. In COVID-19 and other acute viral pneumonias, persistent inflammatory changes and fibrotic patterns are observed in follow-up CT scans in 40-56% patients^2^. In cystic fibrosis (CF), neutrophilic inflammation drives progressive bronchiectasis from early childhood that often remains in small airway regions^3^. Although these diseases differ in etiology, airway remodeling represents a convergent pathological process with important prognostic significance^4^.

Airway remodeling typically begins in small, distal airways^4^ whose dimensions approach the CT resolution. Visual assessment of conducting airways on CT remains challenging due to branch orientation, partial-volume effects, and regional disease heterogeneity. Moreover, airway remodeling is not spatially uniform: idiopathic pulmonary fibrosis (IPF) has a predilection for the lower lobes^5^, COVID-19 showed lower lobar involvement in over 85% patients in a small retrospective study^6^, and early bronchiectasis in CF may be confined to a single lobe^7^. These structural abnormalities can precede overt symptoms or measurable pulmonary function impairment^8,9^, allowing potentially irreversible airway injury to accumulate before becoming clinically apparent.

Existing quantitative tools including AirQuant^10^, AirMorph^11^, and commercial packages^12,13^ have been validated in various lung diseases, but capture only a subset of the structural airway alterations through a small set of predefined descriptors. For instance, while inter-branch tapering is designed to measure cylindrical bronchiectasis^14^, bronchiectasis may begin with irregular mucus deposition, wall thickening, and mucus plugging before progressing to cylindrical, varicose, or cystic dilation^15,16^. Similarly, fibrosis can distort airway path geometry and branching architecture, while persistent inflammation may alter airway topology and tree complexity^17^. These changes represent distinct biological axes of injury, and a broader feature set could better support measurement of known disease manifestations and discovery of previously unrecognized airway phenotypes.

We introduce Radiomics of the Airway (RadAr), an automated pipeline that, given a CT scan, segments the airway, labels the bronchial hierarchy and lobes, and extracts a comprehensive suite of interpretable airway features spanning individual branches to the entire airway tree. RadAr quantifies complementary dimensions of airway deformation including luminal dimensions, novel analytic measures of tapering and diameter-irregularity, three-dimensional path geometry and branching distortion, and summarizes overall airway complexity using global tree descriptors. We evaluated whether these measurements could identify disease-specific and cross-disease structural phenotypes associated with clinically relevant outcomes, pulmonary physiology, and airway inflammation across fibrotic interstitial lung disease, COVID-19, progressive pulmonary fibrosis, and pediatric cystic fibrosis. We additionally built an interactive, browser-based portal that runs the full pipeline with a public browse-only demonstration hosted at https://pushkarmutha.github.io/RadAr-Demo.

## Materials and Methods

### Dataset

#### Fibrotic Interstitial Lung Disease

This retrospective study utilized CT scans from N=147 (D_1_) fILD patients from the public AIIB23 challenge^18^. This cohort included patients with IPF (36%), other ILDs (57·6%), and other respiratory conditions (6·4%). The data was divided into a balanced training set (D_1_^train^, N=68) by undersampling the majority class, and a holdout challenge validation set (D_1_^test^, N=52). Mortality outcome labels for D_1_^test^ were withheld, and model predictions were submitted to the challenge evaluation portal for scoring against ground truth.

#### COVID-19

Non-contrast CT scans were obtained from two publicly available datasets. The STOIC2021-COVID-19 dataset^19^ included N=1205 COVID-19 patients from 20 French university hospitals. CTs with imaging artifacts, an opaque hemithorax, or complete airway segmentation failure were excluded, yielding N=1164 patients (884 mild, 280 severe). Severe COVID-19 was defined as the need for mechanical ventilation, intubation, or death. The cohort was divided into a majority-class undersampled balanced training set (D_2_^train^), (N=200; 100 severe), and a holdout testing set (D_2_^test^) with remaining patients (N=964; 180 severe). The Stony Brook University (SBU) COVID-19 dataset^20^ (D_2_^ext^, N = 205) was used for external validation. After applying the same exclusion criteria, N=189 patients (63 severe) were retained. Patients with undetectable lobar airways were excluded from lobe-level analyses.

#### Progressive Pulmonary Fibrosis

Deidentified CT scans from N = 9 non-idiopathic PPF patients (D_3_) were acquired at Duke University as part of the Xenon ILD study (NCT05241275). Participants also underwent pulmonary function testing (PFT) and hyperpolarized ¹²⁹Xe magnetic resonance imaging (MRI). MRI-derived regional gas-exchange metrics were obtained for ventilation, membrane uptake, and red blood cell transfer compartments. Each compartment was further stratified into defect, low, and high subregions based on voxel-wise signal intensity distributions.

#### Pediatric Cystic Fibrosis

A pilot cohort of 11 children (8 males) with CF (D_4_) at two years of age was obtained from the Integrated Monitoring Platform for Early Disease Events in Cystic Fibrosis study at Children’s Healthcare of Atlanta^21^. Each child underwent same-day ultra-low-dose inspiratory and expiratory CT, bronchoalveolar lavage (BAL) sampling, induced sputum and blood sampling. BAL collections were performed using normal saline aliquots (1 mL/kg per lobe), and the collected samples were analyzed using previously published protocols^21–23^. Five children had at least one prior pulmonary exacerbation (PEx), defined as the need for intravenous or oral antibiotics.

### RadAr Pipeline

Airways were segmented using two publicly available models, NaviAirway^24^ and Fuzzy Attention Neural Network^25^ and their outputs were merged (Figure 1A). A three-dimensional centerline skeleton of the airway tree (Figure 1B) was then extracted from the airway mask.

**Figure 1:**
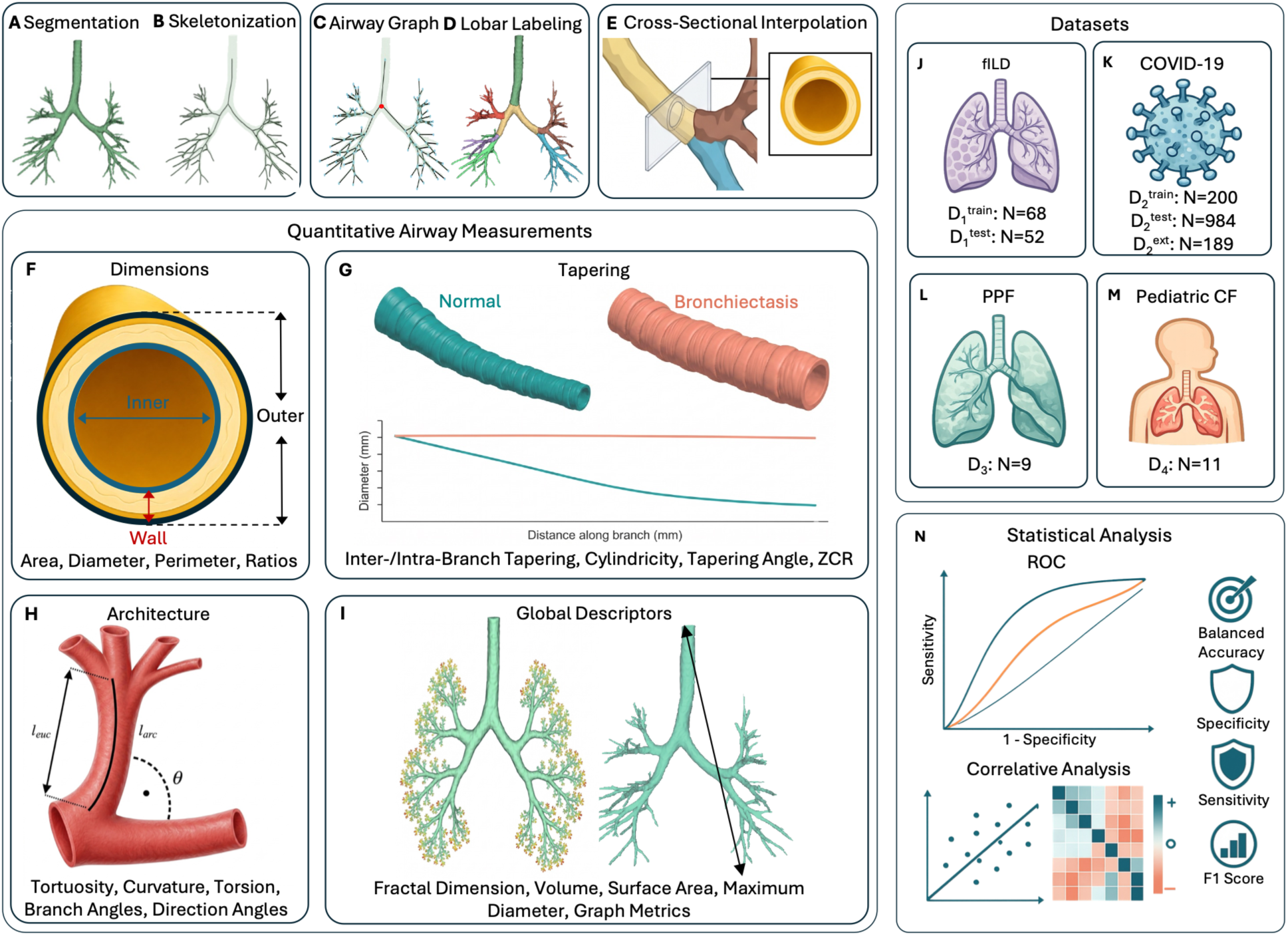
Overview of the RadAr airway phenotyping pipeline and study design. (A-E) Pipeline stages include: (A) airway segmentation from CT, (B) skeletonization, (C) airway graph construction with carina identification, (D) automated anatomy-aware lobar labeling, and (E) extraction of cross-sectional planes perpendicular to the local branch tangent for luminal and wall measurement. (F-I) Quantitative airway measurements: (F) cross-sectional dimensions; (G) tapering features quantifying the loss of normal distal narrowing in bronchiectasis; (H) architectural features capturing three-dimensional path geometry, and (I) global tree descriptors. (J-M) Evaluation cohorts: (J) fibrotic ILD; (K) COVID-19; (L) progressive pulmonary fibrosis; and (M) pediatric cystic fibrosis. (N) Statistical analysis comprised of area under the receiver operating characteristic curve, balanced accuracy, sensitivity, specificity, and F1 score, and correlative analysis between airway features and clinical, functional, and inflammatory measures. Panels E-N created using AI (ChatGPT 5, OpenAI). ZCR: Zero Crossing Rate.

### Airway Graph Construction and Carina Identification

The skeleton was parsed to extract individual branch paths, which were assembled into a directed acyclic graph (DAG), with the superior-most skeleton point designated as the tracheal root. The carina was identified as the DAG node with highest out-closeness centrality (Figure 1C). The two immediate descendants of the carina were assigned as the left and right main bronchi based on their mediolateral position relative to the tracheal midline, and airway generations were assigned recursively beginning from generation 1 at the main bronchi.

### Automated Lobar Labelling

Lobar assignments were determined using an anatomy-aware rule-based algorithm (Figure 1D) adapted from previously described methods^10,26^. On the left side, generation-2 branches were classified as the left upper lobe (LUL) or left lower lobe (LLL) based on the axial coordinate of their most distal terminal nodes. The left middle lobe (LML) was identified among the generation-3 LUL branches as the most distal inferior descendant. On the right side, the right upper lobe (RUL) and right bronchus intermedius (RBI) were separated at generation 2 using endpoint axial coordinates, while the right middle and lower lobes (RML and RLL, respectively) were distinguished within RBI branches based on the geometric relationship between leaf-node axial and coronal coordinates. Lobar labels were propagated recursively to all descendant branches. A quality control step was performed to semi-automatically correct any lobe labelling errors (Supplement S2.6).

### Airway Lumen and Wall Measurement

Airway branch coordinates were smoothed and resampled at 0·5 mm intervals. At each sampling point, a cross-sectional plane perpendicular to the local tangent vector was extracted (Figure 1E). Lumen and wall boundaries within each cross-section were estimated using a modified airway-segmentation-guided ray-casting based full-width-at-half-maximum method^10,27^. Ellipses were fitted independently to the inner and outer boundary point sets to compute airway lumen and wall geometry (Figure 1F).

### Feature Extraction

Four complementary categories of airway features were computed to quantify airway morphology across multiple spatial scales.

#### Airway dimensions

For each cross-section along an airway branch, inner, outer and wall measurements of area, diameter, perimeter, eccentricity, elongation, and their corresponding ratios were computed (Figure 1F). Pi10 was computed by regressing the square root of wall area against inner perimeter and extracting the predicted value at an inner perimeter of 10 mm^28^.

#### Tapering

Inner lumen diameter was modelled as a linear function of cumulative arc-length distance from the branch origin (Figure 1G) using Huber robust regression^29^. Novel quantitative measures of tapering and cylindricity were computed for all airway branches using the regression slope and intercept. Closed-form measurements of average diameter, diameter standard deviation, intra- and inter-branch tapering, taper rate, tapering angle, and cylindricity, branch volume, branch surface area, branch surface area-to-volume (SA:V) ratio were derived in closed form. Because pre-bronchiectatic changes involve irregular mucus deposition along the airway wall, the number of zero-crossings and the zero-crossing rate of regression residuals were used to quantify diameter irregularity along each branch.

#### Architectural distortion

Branch tortuosity was computed as the ratio of arc length to chord length. Curvature, torsion, and direction angles^30^ were extracted at points along each airway branch. Branching angles were derived from the dot product of branch direction vectors. (Figure 1H). First-order statistics were computed across all points within each branch for curvature, torsion, and direction angle.

#### Global airway descriptors

Nine graph-theoretic features were extracted from the DAG for each region. Fractal dimension, total airway count, and voxel-based volumetric features were also computed. Carina-to-terminal-endpoint path lengths were computed for each lobe and served as a measure of tree asymmetry (Figure 1I).

#### Aggregate feature vectors

Branch-level features were summarized at the patient level by computing eight summary statistics for each feature. Aggregation was performed separately for the entire airway tree (excluding trachea and main bronchi) and for each of six lobar regions, yielding 462 features per patient for the overall airway and for each lobe. A detailed description and analytical equations of the RadAr pipeline are provided in Supplement S2.

### Interactive Web Portal

We implemented a browser-based portal that executes the full pipeline on an uploaded CT. The interface renders the three-dimensional airway tree and maps the extracted per-branch features onto the tree as a color-coded heatmap for spatial interpretation. A relabeling view allows a user to reassign the lobe of any branch to correct automated labelling errors. All components of the RadAr pipeline and subsequent data analyses were implemented manually, only the interactive web portal was developed with the assistance of an AI coding tool (Claude Code, Anthropic).

### Statistical Analysis

Overall airway and lobe-specific classification models were trained to predict 63-week mortality in fILD using D_1_^train^ and COVID-19 severity using D_2_^train^. Following test-retest repeatability analysis (Supplement S3), highly correlated features were removed, informative features were selected, z-score standardized, and logistic regression classifiers were trained within a cross-validation framework. Model performance was evaluated using area under the receiver operating characteristic curve (AUC), average precision (AP), balanced accuracy, sensitivity, specificity, and F1 score. To assess the independent value of RadAr for predicting COVID-19 severity, multivariable logistic regression models were fitted on D_2_^ext^ adjusting for systolic blood pressure, respiratory rate, heart rate, pulse oximetry below 90% (binary), and age. Odds ratios, 95% confidence intervals, and p-values were reported. Equivalent adjustment was not possible for D_2_ or D_1_ since clinical information was unavailable. Additional details on classifier training and evaluation are provided in Supplement S4.

To identify airway phenotypes shared across diseases, overall-airway features from D_1_, D_2_ and D_2_^ext^ cohorts were pooled. Repeatable features were retained, features were clipped at the 1^st^-99^th^ percentiles, z-score standardized within each dataset, and highly correlated features (|r|>0·90) were removed. Consensus clustering was performed for k=2-8 using 250 resamples of 75% of patients and Ward agglomerative clustering^31^. The optimal k was selected from the consensus cumulative distribution function, and final phenotypes were assigned by k-means clustering of the consensus matrix. Phenotypes were ordered by increasing outcome rate. Outcomes were used only for phenotype ordering and characterization; the analysis was unsupervised and hypothesis-generating.

For PPF, RadAr features from the overall airway tree were grouped into five morphological categories: luminal dimensions, branch length, tapering, tortuosity, and global airway-tree descriptors. Two principal components were retained for each category, explaining 57%-89% of within-category variance, and were correlated with PFT and ¹²⁹Xe MRI gas-exchange measures using Spearman correlation. For pediatric CF, eight domain-selected tapering features were correlated with four BAL-derived inflammatory markers from the RML and LML using Spearman correlation. Differences according to prior PEx history were evaluated using the Mann-Whitney U test. Features with p<0·05 were considered significant while those with p<0·10 were considered nominally associated. Given the small sample sizes of the PPF and pediatric CF cohorts, multiple comparison correction was not applied, and all findings from these analyses are considered exploratory and hypothesis-generating.

## Results

We evaluated RadAr measurements across complementary dimensions of imaging-biomarker validity: technical repeatability (Supplement S3), prognostic association, structure-function association, structure-inflammation association, and unsupervised phenotype discovery.

### 63-Week Mortality Prediction in Fibrotic Interstitial Lung Disease

In the fILD cohort, RadAr features associated with 63-week mortality were dominated by architectural distortion metrics. The overall airway model selected tortuosity, branching angle heterogeneity, and curvature (Figure 2A), indicating that global airway path irregularity was the principal morphological signature of mortality risk. Similar feature patterns were observed in the LLL and RML models, whereas the RLL model additionally selected Pi10 and fractal dimension, suggesting complementary contributions from airway wall thickening and global tree complexity.

**Figure 2:**
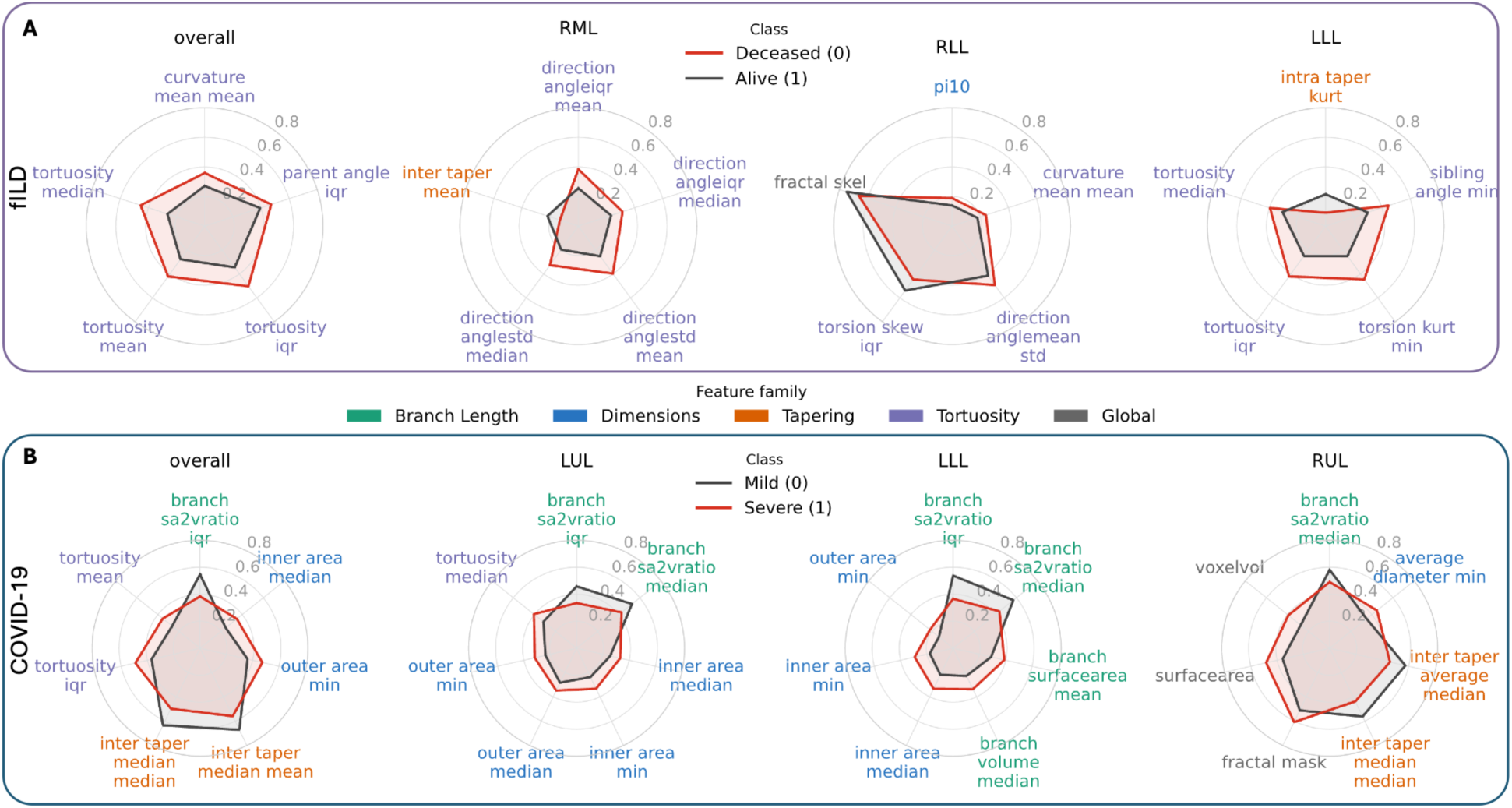
Spiderweb plots showing average values of top selected features scaled between 0 and 1 across (A) deceased (red) and alive (blue) patients in fILD cohort, and (B) mild (blue) vs. severe (red) COVID-19 patients. Individual feature names are colored according to the feature family. RUL: Right Upper Lobe, RML: Right Midde Lobe, RLL: Right Lower Lobe, LUL: Left Upper Lobe, LLL: Left Lower Lobe.

These RadAr signatures were prognostic in a blinded validation setting. The overall airway model achieved a balanced accuracy of 0·654 (sensitivity 0·615, specificity 0·692, F1 = 0·64) on D_1_^test^. Lobar model performance ranged from 0·519 to 0·635% balanced accuracy. With the RLL and RML models achieving 0·635, and the LLL model achieving 0·596. In contrast, upper-lobe models showed limited discriminative value, particularly the RUL model, which performed near chance (Figure 3A). Together, these findings suggest that mortality-associated airway remodeling in ILD is preferentially captured within the lower and middle lung regions, consistent with the regional distribution of fibrotic remodeling.

**Figure 3:**
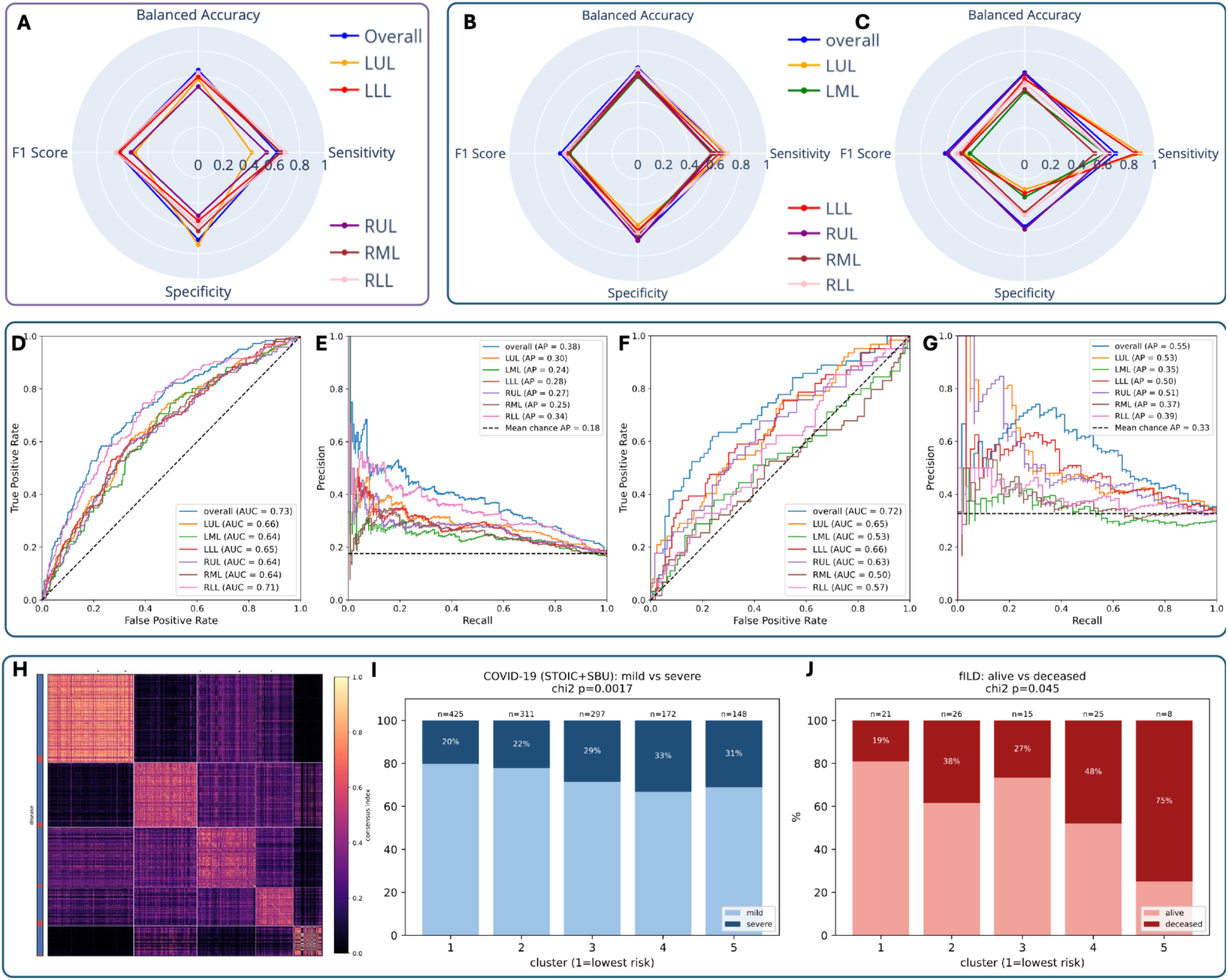
RadAr performance for fILD mortality and COVID-19 severity prediction. Spiderweb plots with classification metrics for (A) 63-week mortality prediction in fILD (D_1_^test^); COVID-19 severity prediction in the (B) internal holdout cohort (D_2_^test^), and (C) external validation cohort (D_2_^ext^); AUC and AUPRC plots for (D-E) internal holdout cohort (D_2_^test^) and (F-G) external validation cohort (D_2_^ext^). (H) Consensus heatmap with five clusters, and illustration of proportion of (I) severe COVID-19 and (J) deceased patients from the fILD cohort across the five airway phenotypes.fILD: Fibrotic Interstitial Lung Disease, RUL: Right Upper Lobe, RML: Right Midde Lobe, RLL: Right Lower Lobe, LUL: Left Upper Lobe, LML: Left Middle Lobe, LLL: Left Lower Lobe, AP: Average Precision.

### COVID-19 Severity Prediction

The airway signature of severe COVID-19 was characterized by convergent selection of branch SA:V ratio, lumen area, and tortuosity across the overall and lobar models (Figure 2B). Reduction in branch SA:V ratio was driven by increased branch volume relative to branch SA or airway count in severe COVID-19 (Figure S10A-B). Together, these features suggest a morphology of airway dilation, loss of normal tapering, and increased path distortion. The recurrence of these features across independently trained models supports a lobe-independent airway phenotype associated with severe acute disease. The RLL model additionally selected curvature features, consistent with localized airway-path distortion in regions commonly affected by COVID-19 pneumonia.

The overall airway model demonstrated consistent discrimination between mild and severe COVID-19 across both internal and external evaluation sets, with AUCs of 0·725, AP of 0·38 (chance: 0·187) on D_2_^test^ and AUC of 0·719, AP of 0·549 (chance: 0·333) on D_2_^ext^. Lobar performance was more heterogeneous, with AUCs ranging from 0·636 to 0·713 on D_2_^test^ (Figure 3B,D,E) and from 0·496 to 0·656 on D_2_^ext^ (Figure 3C,F,G). Left-sided lobes and the RUL model generalized more consistently, whereas RML, RLL, and LML models demonstrated limited external generalization. The overall airway prediction remained independently associated with severe COVID-19 after adjustment for clinical variables (OR = 2·32, 95% CI 1·16-4·66, p = 0·017), with similar associations observed for the LUL, RUL, and LLL lobar models (Figure S9). Across all adjusted models, oxygen saturation below 90% remained the strongest clinical predictor of severity (OR range: 3·06 -4·50, all p < 0·01).

### Airway Phenotypes Across fILD and COVID-19

Consensus clustering identified five reproducible airway phenotypes (P1-P5) that were independent of diagnosis, with fILD and COVID-19 patients represented across all phenotypes. The phenotypes formed a structural continuum from extensive, finely branched, thin-walled airway trees (P1) to sparse, dilated, thick-walled, and tortuous trees (P5), with P2-P4 representing intermediate states (Figure 3H-J, Figure S11, Figure S12). This continuum was associated with outcomes in both diseases (Table S2). From P1 to P5, severe COVID-19 increased from 20% to 33% (<0·0001), and fILD mortality increased from 19% to 75% (p=0·008). The phenotype axis captured the same features identified by supervised models of luminal dilation, reduced branch SA:V ratio, tortuosity, and wall thickening, despite being derived without outcome labels.

### Structure-Function Association in Progressive Pulmonary Fibrosis

In PPF, airway morphology was most consistently associated with spirometric and volumetric measures. Branch length principal component 1 (PC1), reflecting greater airway length and volume, demonstrated the strongest associations with preserved lung function, including FVC (ρ = 0·83, p = 0·005), FEF25-75 (ρ = 0·87, p = 0·002), and alveolar volume (ρ = 0·77, p = 0·016). Tapering PC1 similarly correlated with FVC (ρ = 0·78, p = 0·013), FEV1 (ρ = 0·77, p = 0·016), and alveolar volume (ρ = 0·73, p = 0·025), indicating that preservation of airway tapering tracks with better global lung function (Figure 4).

**Figure 4:**
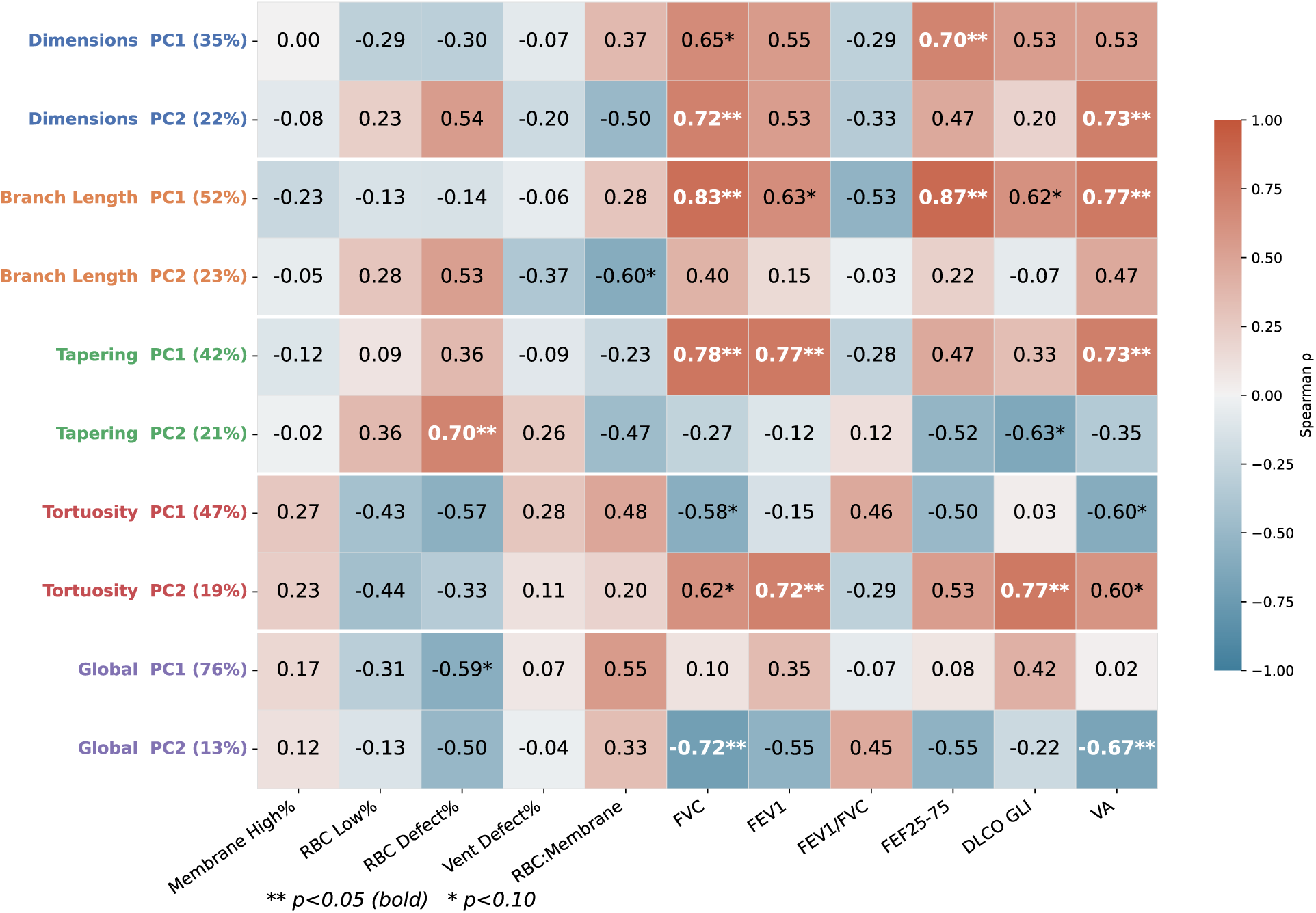
Structure-function associations in progressive pulmonary fibrosis. Spearman rank correlations between principal components from five airway feature categories (dimensions, branch length, tapering, tortuosity, global tree descriptors; within-category variance explained shown in parentheses) and ¹²⁹Xe MRI gas exchange metrics and pulmonary function indices. Bold values indicate p < 0·05 (**); starred values, p < 0·10 (*). PC: Principal Component, RBC: Red Blood Cell, Vent.: Ventilation, FVC: Forced Vital Capacity, FEV1: Forced Expiratory Volume in one second, FEF25-75: Forced Expiratory Flow between 25% and 75% of vital capacity, DLCO: Diffusing Capacity of the Lungs for Carbon Monoxide, GLI: Global Lung Function Initiative, VA: Alveolar Volume.

Other feature families captured complementary structural axes. Airway-dimension components were associated with FEF25-75, FVC, and alveolar volume, while tortuosity PC2 correlated with FEV1 (ρ = 0·72, p = 0·030) and DLCO (ρ = 0·77, p = 0·016). Global airway-tree descriptors were negatively associated with FVC (ρ = −0·72, p = 0·030) and alveolar volume (ρ = −0·67, p = 0·050), consistent with global airway remodeling as a marker of more advanced disease. Among ¹²⁹Xe

MRI metrics, only tapering PC2 correlated with RBC Defect% (ρ = 0·70, p = 0·035), suggesting that RadAr primarily captures structural remodeling and lung-volume impairment rather than alveolar-capillary gas-transfer abnormalities.

### Early Airway Remodeling in Pediatric Cystic Fibrosis

Reduced airway tapering was consistently associated with early neutrophilic airway inflammation. Significant inverse correlations were observed between median intra-branch tapering and neutrophil percentage in BAL fluid from both RML (ρ = −0·64, p < 0·05) and LML (ρ = −0·78, p < 0·05) (Figure 5A). Children with a history of prior PEx demonstrated significantly reduced intra-branch tapering and tapering angle, together with increased cylindricity (all p<0·05) compared to those without prior PEx (Figure 5B). These findings suggest that loss of airway tapering accompanies early neutrophilic inflammation and clinical disease activity in young children with CF. Other airway features demonstrated variable, non-significant associations with BAL inflammatory markers, likely reflecting limited sample size.

**Figure 5:**
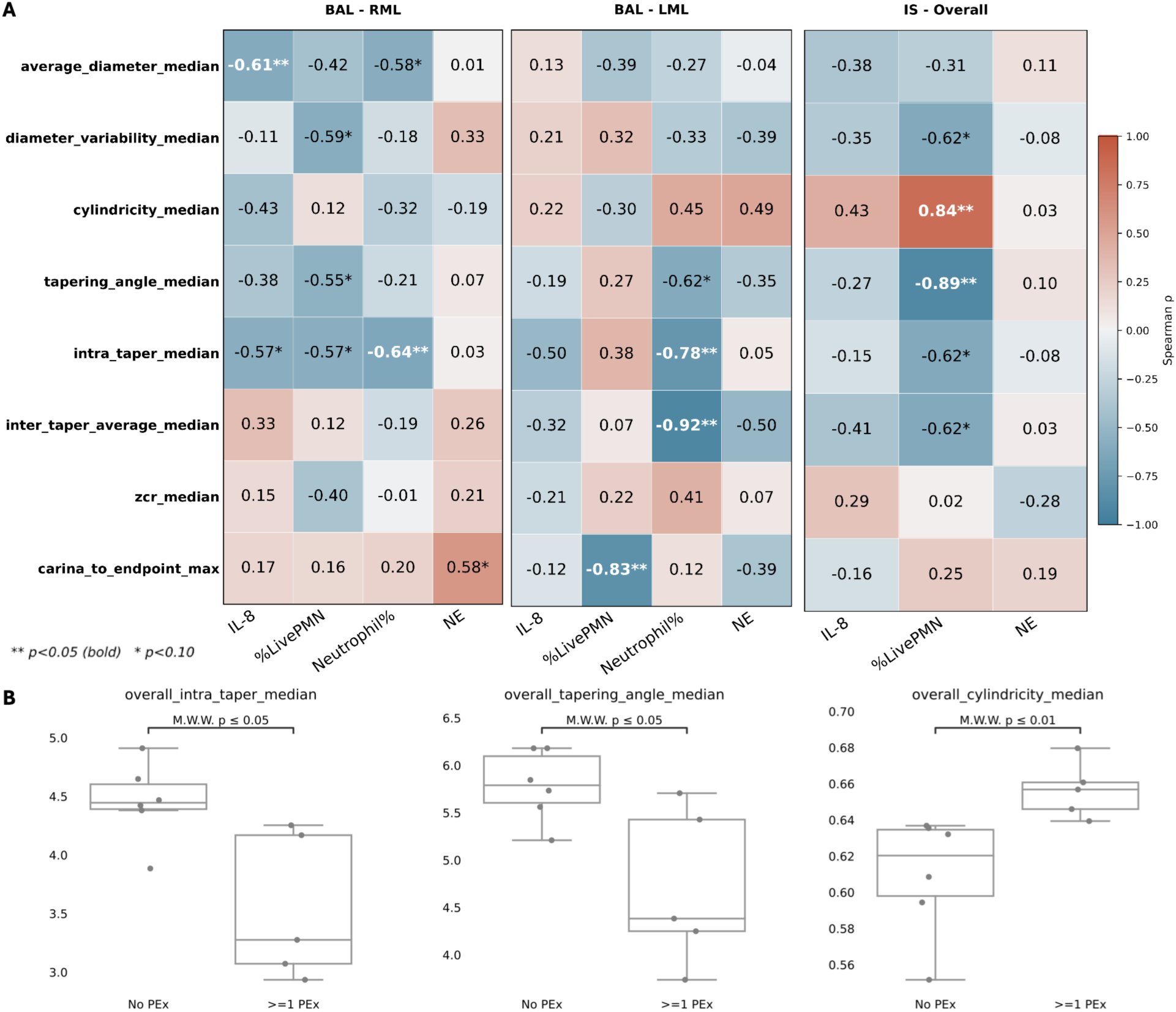
Airway structure, inflammation, and exacerbation history in pediatric cystic fibrosis. (A) Spearman correlations between RadAr airway features and inflammatory markers (IL-8, %LivePMN, neutrophil percentage, neutrophil elastase) measured in bronchoalveolar lavage from the RML and LML, and in induced sputum. Bold values indicate p < 0·05 (**); starred values, p < 0·10 (*). (B) Overall intra-branch tapering, tapering angle, and cylindricity median in children with no prior pulmonary exacerbations (No PEx) versus ≥ 1 PEx. BAL: Broncho-alveolar Lavage, IS: Induced Sputum, RML: Right Middle Lobe, LML: Left Middle Lobe, IL-8: Interlerukin-8, PMN: Polymorphonuclear cells, NE: Neutrophil Elastase, M.W.W: Mann-Whitney-Wilcoxon.

**Figure 6:**
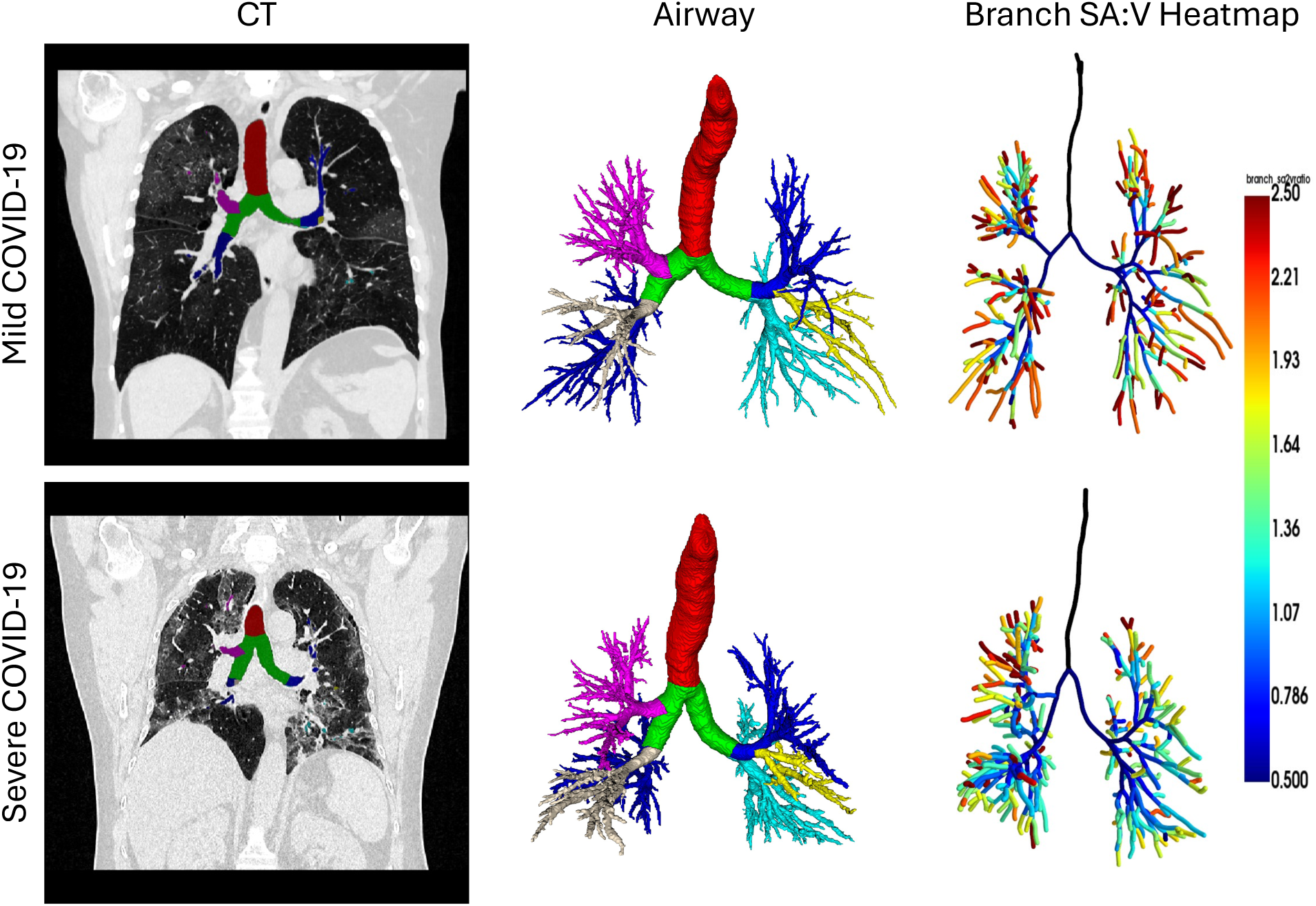
Qualitative visualization of CT scans, associated lobe labelled airways, and branch-wise surface area to volume (SA:V) ratio feature heatmap in airway volume and airway count matched mild (top) and severe (bottom) COVID-19 patient.

## Discussion

Across a spectrum of etiologies spanning restrictive, obstructive, and mixed lung diseases, RadAr recovered structural signals consistent with each disease’s underlying pathology. In fILD, a restrictive disease, the prognostic features were dominated by architectural distortion and concentrated in the lower lobes, consistent with the basal, peripheral distribution of IPF-pattern fibrosis and with traction bronchiectasis associated with peribronchial fibrosis^5^. In COVID-19, where physiology is more heterogeneous and can combine restrictive and obstructive elements, the top features were dominated by luminal caliber, and the resulting score remained associated with severity after adjustment for clinico-demographic variables indicating that it captured structural information beyond routine clinical markers. In PPF, airway features correlated with spirometric and volumetric indices consistent with conducting-airway geometry indexing the volumetric, restrictive axis of disease^32,33^. The pediatric CF cohort, representing an obstructive, bronchiectasis-predominant phenotype, was evaluated prior to the initiation of CFTR modulator therapy and showed reduced tapering and increased cylindricity was associated with greater neutrophilic inflammation and with a history of pulmonary exacerbation, recapitulating the established link between airway neutrophilic inflammation and the development of bronchiectasis in early CF^3,9^.

Interestingly, severe COVID-19 was associated with a reduced median and inter-quartile range of per-branch SA:V ratio, both overall and across individual lobes. This is directionally consistent with the findings of Bodduluri et al.^34^, who reported higher all-cause mortality with reduced SA:V ratio in chronic obstructive pulmonary disease (COPD). However, they computed a single, voxel-based SA:V ratio over the entire airway tree, susceptible to partial volume effects and segmentation noise. RadAr instead analytically derived robust estimates of SA:V ratio for each branch, providing a more granular and sensitive assessment of changes in SA:V ratio with severe disease. Accordingly, the median inner lumen area was also higher in severe patients. This interpretation is biologically consistent with the acute pathophysiology of COVID-19, in which inflammation and edema alter airway caliber to produce a frequently reversible bronchial dilatation in contrast to the fixed distortion of established fibrosis^35^. Beyond disease-specific models, unsupervised analysis of the pooled fILD and COVID-19 cohorts identified a shared continuum of airway remodeling that was prognostic in both diseases. The highest-risk phenotype characterized by sparse, dilated, thick-walled, and tortuous airways identified features selected by supervised models for severe COVID-19 and fILD mortality, despite being derived without outcome labels.

RadAr’s multi-scale feature set can be extended to diseases where airway remodeling may be implicated but has not been quantitatively characterized. For example, while acute COVID-19 is no longer a major clinical burden, RadAr features could be repurposed to build a signature for identifying patients at risk of developing long-COVID syndrome^36^ or to track treatment response as therapies for long-COVID mature. In CF, CFTR modulators have restored CFTR function to near-normal levels and helped modify the natural history of lung disease progression, with case reports of bronchiectasis reversal on serial CTs in treated patients^37^. RadAr could detect early airway damage not routinely quantified on visual CT inspection in young children who would benefit from additional preventative interventions before bronchiectasis becomes irreversible. More broadly, conventional trial endpoints for pulmonary therapies such as spirometry or PEx rate are insensitive to early structural disease and regional heterogeneity^38^. Direct quantification of tapering loss, cylindricity or architectural distortion could serve as imaging-based surrogate endpoints that more precisely capture the effects of therapy. Beyond primary pulmonary diseases, airway remodeling is increasingly recognized as a contributor to or consequence of systemic disorders and their treatments. For instance, checkpoint-inhibitor or radiation induced pneumonitis have shown airway architectural consequences^39,40^. RadAr extracts a broad set of airway attributes, and which can be assembled into disease-specific biomarkers or composite signatures for assessing the impact of therapy, exposure, or comorbidity; building diagnostic, prognostic, or treatment-response signatures from retrospective imaging data; and, with sufficient validation, deployed in prospective clinical trials as outcome measures.

Recent work has applied deep learning to airway quantification in pulmonary fibrosis with a prognostic intent, but each effort has been confined to a single disease and to a small number of hand-selected measurements. Deep-learning quantification of total airway volume predicted mortality independently of total fibrosis extent in the Australian IPF registry^41^, and the recently described SABRE engine derived explainable airway biomarkers that prognosticated independently of lung function in a national registry of more than 1700 patients^42^. RadAr differs in three respects: 1) it computes a wide set of biologically interpretable features; 2) it resolves these features by lobe, reflecting the regional heterogeneity of airway remodeling; and 3) it has been evaluated across four diseases that span the acute and chronic, the obstructive and restrictive types, and the adult and pediatric settings.

Several limitations of our study warrant discussion. First, the PPF and CF analyses used small cohorts, without multiple-comparison correction, and are hypothesis-generating requiring replication in larger, prospectively enrolled samples. Second, performance was heterogeneous across lobes in COVID-19, with middle-lobe models failing to generalize to the external cohort, likely reflecting the smaller caliber and greater anatomic variability of these regions, where airway segmentation and cross-sectional measurement are less reliable. Third, CT acquisition parameter impact was not explicitly evaluated, as the de-identified public cohorts lacked acquisition metadata and scanner parameters. However, the robust performance in the independent SBU external validation set, drawn from a different continent, as well as the test-retest stability analysis demonstrates the robustness of RadAr features to acquisition setting. Finally, all analyses were cross-sectional and retrospective, and the value of these features for tracking disease progression and treatment response remains to be demonstrated longitudinally and prospectively.

In conclusion, RadAr provides an automated, lobe-resolved, and interpretable characterization of airway morphology across fILD, COVID-19, PPF, and pediatric CF from routinely acquired chest CT. Since RadAr provides objective measurements that map onto concepts radiologists already reason with, such as loss of tapering, tortuosity, and wall thickening, this framework can be extended into other lung diseases impacting airways. Prospective and longitudinal validation, together with integration of airway phenotypes with parenchymal, vascular, and functional imaging, can establish these measurements as novel surrogate endpoints in pulmonary trials as well as clinically actionable biomarkers of airway remodeling.

## Ethics Statement

This retrospective study included both institutional and publicly available datasets. The institutional progressive pulmonary fibrosis and cystic fibrosis cohorts were acquired under protocols approved by the Institutional Review Boards (IRB), and written informed consent was obtained from all participants. IRB of Emory University and Duke University gave ethical approval for this work under protocol IRB00097352 and Pro00109322, respectively. The remaining datasets were obtained from publicly available repositories and were used in accordance with their respective data use agreements and institutional policies.

## Data Availability Statement

The COVID-19 datasets analyzed in this study are publicly available. The STOIC2021 COVID-19 cohort can be accessed through the STOIC2021 COVID-19 AI Challenge (https://stoic2021.grand-challenge.org), and the Stony Brook University COVID-19 cohort is available from The Cancer Imaging Archive (https://www.cancerimagingarchive.net/collection/covid-19-ny-sbu/). The fibrotic interstitial lung disease cohort was obtained from the AIIB23 challenge (https://codalab.lisn.upsaclay.fr/competitions/13256). All public datasets were used in accordance with their respective data use agreements. The progressive pulmonary fibrosis and pediatric cystic fibrosis datasets are institutional and are not publicly available owing to privacy and ethical restrictions, but de-identified data may be available from the corresponding author on reasonable request and subject to institutional review board approval and a data use agreement.

## Acknowledgements

Research reported in this publication was supported by the National Cancer Institute under award numbers -R01CA249992 -01A1, R01CA257612 -01A1, R01CA264017 -01, R01CA268287 -01A1, U01CA113913 -16A1, U01CA269181 -01, U24CA274494-01; U54CA302465-01; R01CA268207-01; the National Heart, Lung and Blood Institute under award numbers -R01HL158071 -01A1; the National Institute of Allergy and Infectious Diseases under award number -R01AI175555 -01A1 and R01HL151277 -01A1, the National Institute of Biomedical Imaging and Bioengineering under award number 75N92022D00015; the National Library of Medicine under award number -R01LM013864 -01A1; the National Institute on Aging under award number -R01AG089759; the National Institute of Diabetes and Digestive and Kidney Diseases under award number -R01DK118431; the Office of the Director, National Institutes of Health under award number 1OT2OD038065-01; the Kidney Mapping and Atlas Project (KMAP) under award number -U01DK133090 -01; the United States Department of Veterans Affairs VA Merit Review award under award number -IBX004121; the VA Biomedical Laboratory Research and Development Service under award numbers -IK6BX006185; the VA Research and Development Office through the Lung Precision Oncology Program -LPOP-L0021; and sponsored research agreements from Astrazeneca, Bristol Myers Squibb, the Prevent Cancer Foundation, Innovation in Cancer Informatics, the Institute for Technology in Health Care, the Breast Cancer Research Foundation and the Scott R. Mackenzie Foundation. The content is solely the responsibility of the authors and does not necessarily represent the official views of the National Institutes of Health, the U.S. Department of Veterans Affairs, or the United States Government.

A U.S. provisional patent application related to the technology described in this manuscript has been filed with the United States Patent and Trademark Office (Application No. 64/099,834, “Systems and Methods for Automated Airway Morphology Quantification,” filed June 26, 2026; with the following named inventors: Anant Madabhushi, Pushkar Mutha, Lokesh Guglani).

## AI and AI-Assisted Technologies Declaration

During the preparation of this work, an AI tool (ChatGPT 5, OpenAI) was used to create Figure 1 panels E-N. Claude Opus 4·8 (Anthropic) was used for grammar and language editing. All components of the RadAr pipeline and subsequent data analyses were implemented manually, only the interactive web portal was developed with the assistance of an AI coding tool (Claude Code, Anthropic). The author reviewed and edited the output as needed and take full responsibility for the content of the published article.

