## Supplementary material for "Radiomics of the Airway (RadAr): Multi-Scale Airway Phenotyping for Disease Characterization on Routine CT Imaging": Data Supplement

**S1: CONOSRT Diagram**

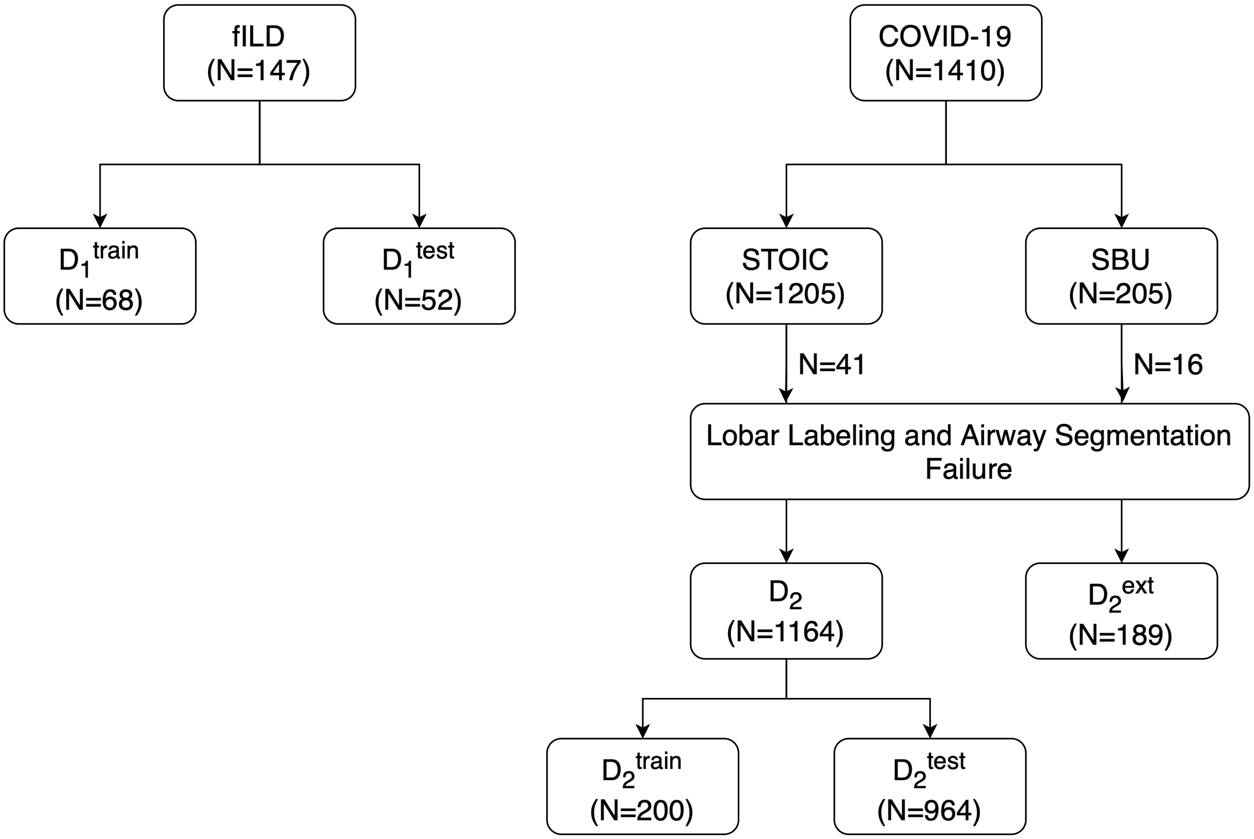

Figure S1: CONSORT flowchart for the fibrotic interstitial lung disease (fILD), and COVID-19 cohorts.

**S2: RadAr Pipeline**

**S2.1: Image Preprocessing**

CT volumes and binary airway segmentations were reoriented to right-anterior-inferior (RAI) convention using SimpleITK's[1–3] DICOMOrient function prior to all processing. A Euclidean Distance Transform (EDT) was computed on the binary airway mask, with anisotropic voxel spacing accounted for, to yield a map of local estimated lumen radii at each airway voxel.

**S2.2: Skeletonization**

A 3D centerline skeleton was extracted using the TEASAR algorithm implemented in Kimimaro[4]. TEASAR parameters were: scale = 1.5, const = 2 mm, pdrf_scale = 100,000, pdrf_exponent = 4, soma_acceptance_threshold = 4 mm, soma_detection_threshold = 750 mm, soma_invalidation_const = 300 mm, soma_invalidation_scale = 2. Post-processing included dust removal (threshold = 4 mm) and tick pruning (threshold = 4 mm). Close disconnected skeleton components were joined. The skeleton was parsed into individual branch paths via junction detection on the binary skeleton image. Terminal branches of ≤ 2 mm were pruned[5].

**S2.3: Airway Graph Construction**

Bifurcation and end points from the skeleton branch were identified using Skan[5]. An undirected graph $G_u$ was assembled with each bifurcation and endpoint as a node. $G_u$ was converted to a directed acyclic graph (DAG) $G$ by performing an edge-directed depth-first search (DFS) from the root node $r_0$.

**Root node selection.** The root $r_0$ was taken as the node whose z-coordinate (superior-inferior axis) was maximum – i.e., the most superior skeleton point, approximating the tracheal apex. When multiple nodes shared the maximum z-coordinate, $r_0$ was selected as the one nearest to the image centroid in the axial plane.

**DAG enforcement.** If G contained cycles (arising from segmentation or skeletonization artifacts), the minimum spanning tree of $G_u$ was computed first, and the DFS was performed on this tree.

**Carina identification.** Closeness centrality $C(v)$ of each node $v$ in the graph $G$ was computed as:

$$C(v) = \frac{n-1}{\sum_{u \neq v} d\,(v,u)}$$

where $d(v,u)$ is the shortest path length from node $v$ to $u$ in $G$ and $n$ is the number of nodes. The carina was defined as the node $c^* = \underset{v}{\operatorname{argmax}}\, C(v)$, reflecting its topological position as the hub from which all downstream airways radiate.

**Airway generation.** Generation was assigned in a breadth-first traversal from $r_0$. The trachea was assigned as generation 0. The two successors of the carina were assigned generation 1 (left and right main bronchi). Each subsequent bifurcation incremented generation by 1 recursively.

#### S2.4: Branch Spline Representation

Raw skeleton path coordinates extracted from the binary skeleton are in voxel space and reflect the discrete, staircase-like path of the skeleton image. At bifurcation nodes, the abrupt directional change between a parent and child branch produces large discontinuities in the finite-difference approximation of the tangent. To obtain smooth, continuously differentiable centerline representations with well-defined tangent vectors, each airway branch underwent spline-based resampling. For each airway branch $j$, its raw skeleton path coordinates were concatenated with those of its immediate parent branch to form an extended coordinate sequence. A 9-point uniform moving-average filter was applied to this concatenated sequence in physical space in millimeters (mm) to suppress discretization noise at bifurcation junctions. A parametric B-spline was then fit to the smoothed extended path. The spline was evaluated and the resulting curve was resampled at uniform 0.5 mm arc-length intervals to yield the final set of sampling points. Unit tangent vectors at each sampling point were obtained from the first derivative of the spline. Because the spline was fit to the extended parent-child sequence, the sampling points span both the parent and the current branch; accordingly, only points from the $j^{th}$ branch's start node onward were retained. This procedure ensured smooth tangent continuity across bifurcations, which is critical for stable perpendicular cross-sectional plane orientation at branch origins. Branches yielding fewer than 4 retained sampling points were flagged as invalid and excluded from feature extraction. The EDT value was interpolated at each sampling point to provide a local radius estimate, and cumulative arc-length distances from the branch origin were computed.

#### S2.5: Automated Lobar Labelling

Lobar assignments were determined by an anatomy-aware rule-based algorithm adapted from previously described methods[6,7].

##### S2.5.1: Left Lung

The generation-1 left airway was labelled as the left main bronchus (LMB). Its two generation-2 children were classified as left upper lobe (LUL) or left lower lobe (LLL) by comparing the axial (z) coordinate of their most distal terminal nodes, found by graph traversal. The child whose most distant descendant has the higher z-coordinate was assigned to LUL; the other to LLL.

A length-ratio correction is applied: let $l_{LLL}$ be the arc length of the detected LLL lobar branch and $l_{LMB}$ that of the left main bronchi (LMB). If $l_{LLL}/l_{LMB} < 0.15$, this is flagged as a likely misclassification due to a short LMB-like stub, and the algorithm re-evaluates the children of its successor.

**Left middle lobe (LML) identification.** Among the generation-3 branches of LUL, the lingula or left middle lobe (LML) is identified as the branch whose most distal terminal node has the lowest z-coordinate (most inferior), and all its descendants are relabeled LML.

#### S2.5.2: Right Lung

The generation-1 right airway was labelled as the right main bronchus (RMB). Its two generation-2 children were classified as right upper lobe (RUL, higher z endpoint) or right bronchus intermedius (RBI, lower z endpoint). A length-ratio check ($l_{RBI}/l_{RMB}$ < 0.15) flags potential RUL misclassification. RBI is re-assigned as generation-1, and its child generations are successively incremented.

**RML and RLL identification.** A subgraph $G_{RBI}$ rooted at the origin of RBI was extracted from the main DAG. All leaf nodes of $G_{RBI}$ were found and the difference between axial and sagittal coordinates (z − y) was computed. The path from the RBI origin to the leaf node with the maximum (z − y) value defined the right middle lobe (RML) trajectory; the path to the minimum (z − y) leaf defined the right lower lobe (RLL). The branch at the first post-RBI bifurcation along the RML path was recursively labelled as RML; all remaining RBI branches were labelled as RLL.

#### S2.5.3: Label Propagation

Lobe labels were propagated from the skeleton to the full airway mask via nearest-neighbor search using a KD-tree. Each airway mask voxel inherited the lobe label of its nearest skeleton voxel.

### S2.6: Quality Control and Interactive Lobar Relabeling

#### S2.6.1: Quality Control Review

A manual quality control (QC) step was performed across all datasets to identify and correct mislabeled airway lobes. QC was conducted by the first author (PM), a graduate researcher in biomedical engineering, with consultation from a pediatric pulmonologist (LG) for challenging or ambiguous cases. Coronal projections of the lobe-labeled airway skeleton were saved automatically for each subject (Figure S2). Each lobe was rendered in a distinct fixed color. All images were reviewed manually and cases with labelling errors were flagged for manual correction or exclusion. Mislabeled airway lobes (Figure S2B) detected during review were corrected using an interactive relabeling mode of the processing pipeline described below. The most common cause of labeling failure was catastrophic or incomplete airway segmentation (Figure S2C), typically arising from extensive parenchymal consolidation, lobar atelectasis, segmentation model failure, or the presence of an endotracheal tube (ETT). Cases with ETT artifact were addressed by manually editing the trachea segmentation to incorporate the ETT as part of the tracheal lumen in ITK-SNAP[8].

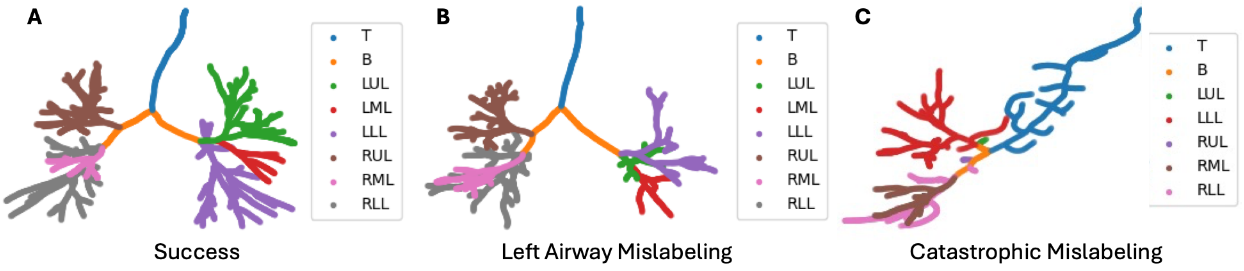

Figure S2: 2D coronal projection of lobe-labelled skeletons for (A) successful lobar labelling, (B) left airway mislabeling where LUL, LML, LLL are incorrectly assigned, (C) catastrophic failure of airway segmentation, skeletonization, and labelling.

#### S2.6.2: Interactive Relabeling GUI

The RadAr pipeline includes a 3D interactive correction workflow after initial automated labelling (Figure S3). A 3D figure is displayed showing the full airway DAG rendered as colored line segments, with each branch colored according to its current automated lobe assignment. The figure is oriented in a coronal anterior view (elevation 0°, azimuth -90°) to match the QC projection image. A radio-button panel on the right side of the figure lists all nine assignable labels: trachea (T), left main bronchus (LMB), right main bronchus (RMB), LUL, LML, LLL, RUL, RML, and RLL. The user interacts with the plot as follows:

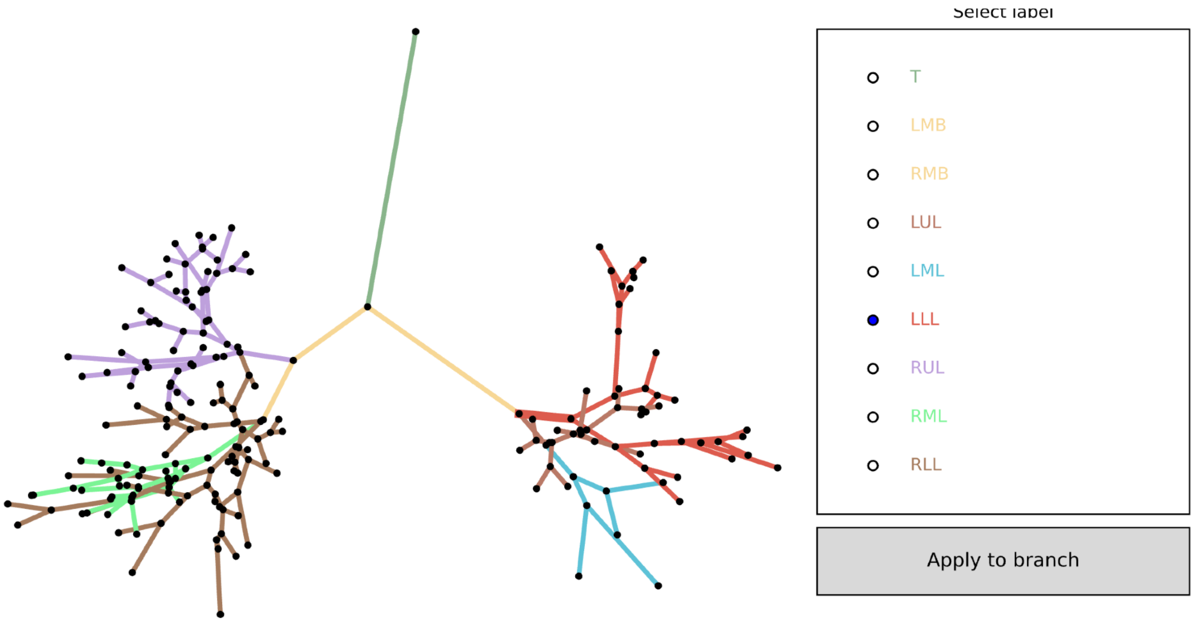

Figure S3: Interactive airway lobe relabeling graphical user interface. The left upper lobar airway branch is highlighted, which auto-populates the radio button incorrectly marked as left lower lobe (LLL).

1. Select a branch. The user clicks on any colored branch segment in the 3D view. The selected branch is highlighted by increasing its line width, and its current lobe label is pre-selected in the radio-button panel.

2. Choose a new label. The user selects the correct lobe label from the radio-button panel.

3. Apply. The user clicks the "Apply to branch" button. The selected label is applied recursively to the clicked branch and all its downstream descendants in the DAG. For LMB and RMB reassignment, a dedicated relabeling routine resets the carina reference point, re-assigns left/right lung identity, and updates generation numbers for the full downstream subtree. After each apply, the graph attributes (lobe assignments, edge weights), lobar metadata, and lobar subgraphs are updated and the corrected labels are immediately saved. Branch colors in the 3D view refresh to reflect the updated assignments.

Steps 1-3 are repeated for each mislabeled region. The GUI remains open until the user closes the figure window.

After relabeling is complete, lobe and lung labels are propagated from the corrected skeleton to the full airway segmentation mask via nearest-neighbor assignment (KD-tree), and processing continues with cross-sectional plane extraction.

**S2.7: Cross-Sectional Plane Extraction**

At the $i^{th}$ sampling point on $j^{th}$ airway branch, $p_{i,j}$, and its tangent $t_{i,j}$, the cross-sectional plane basis vectors were constructed as follows. Let $b_0 \in \{[1,0,0],[0,1,0],[0,0,1]\}$ be the standard basis vector most orthogonal to $t_{i,j}$ (i.e., $\underset{b}{\operatorname{argmax}}\|t_{i,j} \times b\|$). The two in-plane basis vectors are:

$$e_1 = \frac{t_{i,j} \times b_0}{\|t_{i,j} \times b_0\|} \cdot \frac{1}{\Delta s}$$

$$e_2 = \frac{t_{i,j} \times e_1}{\|t_{i,j} \times e_1\|} \cdot \frac{1}{\Delta s}$$

where $\Delta s = (dx, dy, dz)$ is the voxel spacing vector, so that unit steps in $e_1$ and $e_2$ correspond to 1 mm in physical space.

**Grid sampling.** A square patch of size $W \times W$ mm ($W = \max_i(30,\ 4 \times EDT(p_{i,j}) + 10$) at 0.5 mm pixel spacing was placed centered at point $p_{i,j}$. For pixel point $p_{x,y}$ offset from centre $p_{i,j}$, the 3D sample point is:

$$p_{x,y} = p_{i,j} + \left(x - \frac{W}{2\delta}\right)\delta\, e_1 + \left(y - \frac{W}{2\delta}\right)\delta\, \mathrm{e}_2$$

where δ = 0.5 mm. CT values at $p_{xy}$ were interpolated using pre-computed RectBivariateSpline interpolators on each axial slice, with linear weighting between the two nearest slices. Airway mask values were sampled using a RegularGridInterpolator with linear interpolation.

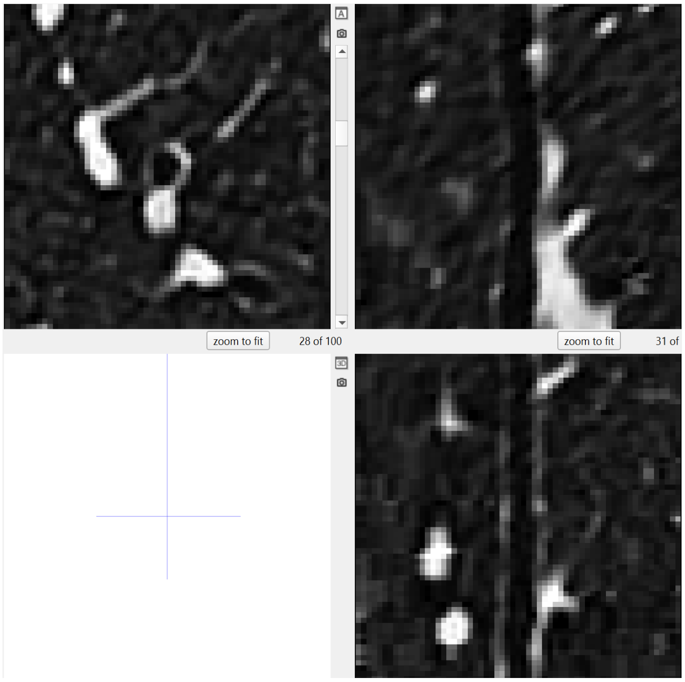

Figure S4: Cross-sectional interpolation of an airway branch.

This produces a stack of cross-sectional image patches for each airway branch (Figure S4). This implementation leveraged Python's broadcasting and vectorization techniques, enabling simultaneous computation of all cross-sectional planes at every point for a single airway branch at once, with significant speed improvements.

#### S2.8: Airway Lumen and Wall Measurement

Lumen and wall boundaries at each cross-section were estimated using a modified airway-segmentation-guided full-width-at-half-maximum (FWHM) method[6,9].

##### S2.8.1: Ray Casting

From the center c of each cross-sectional image (Figure S5A) of shape $(W, W)$, 120 rays were cast at equal angular intervals (Figure S5B). Each ray was sampled at step size 0.1 mm up to a maximum radius of $W/2$ pixels. CT intensity profiles $I_r(n)$ and airway segmentation profiles $S_r(n)$ along each ray were obtained using 5th-order spline interpolation (Figure S5C).

##### S2.8.2: Wall Peak and FWHM Boundary Detection

For each ray r, the lumen-wall transition index $n_{seg}$ was identified as the first position where the segmentation profile $S_r$ fell below 0.5 ($S_r < 0.5$). Among the local maxima of the CT intensity profile $I_r$, the wall peak $w_r$ was selected as the one closest to $n_{seg}$ that lay outside the segmented lumen ($S_r(w_r) < 0.5$). The inner boundary $in_r$ was placed at the half-maximum point between $w_r$ and the nearest local minimum on the lumen side. Symmetrically, the outer boundary $out_r$ was placed at the half-maximum point between $w_r$ and the nearest local minimum on the parenchymal side.

Rays were excluded if the wall peak intensity $I_r(w_r)$ deviated from the median wall peak intensities across all the cast rays by more than 2.24 × median absolute deviation.

The radial co-ordinates of inner and outer boundaries $(in_r, out_r)$ were mapped to the planar co-ordinates yielding inner $(x_{r,in}, y_{r,in})$ and outer $\left(x_{r,out}, y_{r,out}\right)$ boundary points in the image plane, respectively.

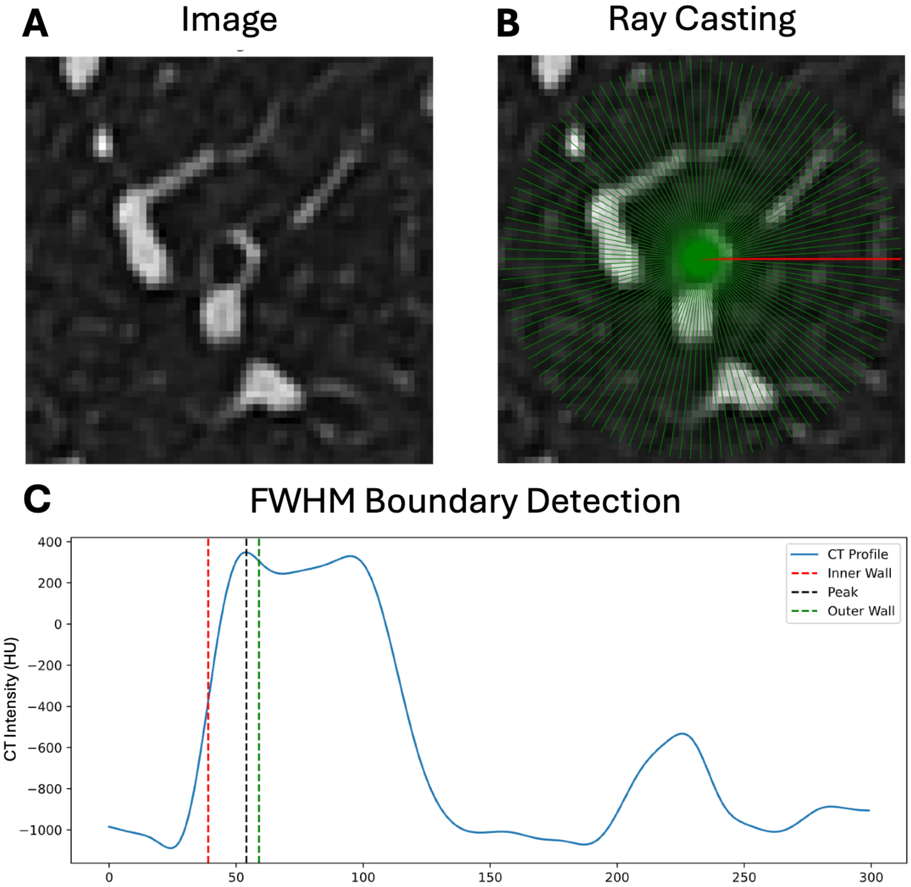

Figure S5: Airway lumen and wall boundary detection using the full-width at half-maximum (FWHM) method for (A) a cross-sectional plane. (B) 120 equidistant rays (green lines) are cast from the center; inner and outer airway boundaries are estimated. The ray highlighted in red's (C) CT intensity profile shows the inner (red), peak (black) and outer (green) intensity locations.

#### S2.8.3 Ellipse Fitting

Inner boundary points $(x_{r,in}, y_{r,in})$ and outer boundary points $\left(x_{r,out}, y_{r,out}\right)$ from all valid rays obtained after outlier removal were fitted with an ellipse, yielding ellipse parameters $(x_c, y_c, a, b, \theta)$ for both inner and outer ellipses (Figure S6). Quantitative measurements were extracted as described below using the ellipse parameters.

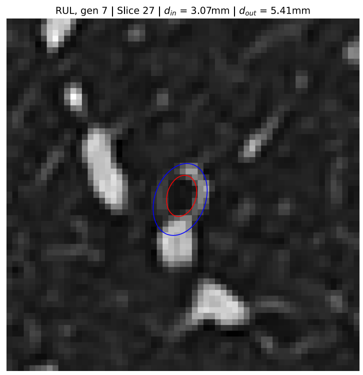

Figure S6: Inner (red) and outer (blue) ellipse fit to FWHM derived boundary points.

**S2.9: Feature Extraction**

**S2.9.1: Cross-Sectional Dimensions**

Cross-sectional measurements of lumen (inner airway), outer airway and airway wall are computed across each point $i$ sampled at equidistant intervals of 0.5mm on branch $j$ in the airway tree.

Let $(a_{in}, b_{in})$ and $(a_{out}, b_{out})$ be the semi-axes $(a \geq b)$ of the inner and outer ellipses, respectively. All linear dimensions are in mm (pixel × 0.5 mm/pixel).

**Areas:** Inner, outer, and wall areas are computed as:

$$A_{in} = \pi\, a_{in} b_{in}, \quad A_{\text{out}} = \pi\, a_{\text{out}} b_{\text{out}}, \quad A_{\text{wall}} = A_{\text{out}} - A_{\text{in}}$$

**Equivalent diameters (area-equivalent circular diameter):** Inner diameter, outer diameter and wall thickness are computed as:

$$d_{\text{in}} = 2\sqrt{A_{\text{in}}/\pi}, \quad d_{\text{out}} = 2\sqrt{A_{\text{out}}/\pi}, \quad WT = \frac{d_{\text{out}} - d_{\text{in}}}{2}$$

**Wall ratios:** Ratios of wall area to outer airway area $(WO)$, inner airway to outer airway area $(IO)$, and wall area to inner airway area (WI) are measured as:

$$WO = A_{\text{wall}}/A_{\text{out}}, \quad IO = A_{\text{in}}/A_{\text{out}}, \quad WI = A_{\text{wall}}/A_{\text{in}}$$

**Perimeters** are computed for inner $(P_{in})$ and outer $(P_{out})$ airway wall using Ramanujan's second approximation:

$$P = \pi(a + b)\left[1 + \frac{3h}{10 + \sqrt{4 - 3h}}\right], \quad h = \frac{(a - b)^2}{(a + b)^2}$$

**Eccentricity and elongation:**

$$e = \sqrt{1 - \text{b}^2/\text{a}^2}, \quad \varepsilon = a/\text{b}$$

Branch level summary for the $j^{th}$ branch is obtained as the median value of these measurements across all valid cross-sections.

**Pi10**: Pi10 is a global airway wall thickness index. For each valid non-tracheal branch j, the square root of wall area was regressed against the inner perimeter using robust Huber regression[10]:

$$\sqrt{A_{\text{wall}}} = \beta_0 + \beta_1 P_{in}$$

Pi10 is the predicted $\sqrt{A_{\text{wall}}}$ at an inner perimeter $(P_{in})$ of 10 mm:

$$\text{Pi10} = \hat{\beta}_0 + \hat{\beta}_1 \times 10$$

**S2.9.2: Tapering Features**

Let the $j^{th}$ airway branch with length $L_j$ have inner equivalent diameter measurements $d_j(l)$ at cumulative arc-length positions $l$ from the beginning of the branch ($l = 0$) to branch endpoint at length $l = L_j$ (in mm). A robust regression model[10] is fit to estimate the inner airway diameter as a function of the distance along an airway branch (Figure S7C).

$$d_j(l) = m_j l + c_j \quad \text{(Eq. 1)}$$

where the slope $m_j$ is the tapering rate and intercept $c_j$ (mm) is a robust estimate of airway lumen diameter ($d_j(0)$) at the branch origin ($l = 0$). Accurate measurements of distal airway branches are non-trivial since (a) their size approaches CT resolution and (b) the contrast between airways and surrounding lung parenchyma is low. The robust estimate of airway diameter from Eq. 1 allows mitigating measurement errors while computing features of branch morphology.

From Eq. 1, closed-form expressions for branch morphology features; branch volume, surface area, branch volume to surface area ratio, average diameter, diameter standard deviation, intra-branch tapering, cylindricity, tapering angle, residual zero-crossings and zero-crossing rate, and inter-branch tapering to quantify the bronchiectasis associated patterns are derived for the $j^{th}$ airway branch.

**Branch Volume ($BV_j$):** Estimating $BV_j$ from voxel data may introduce bias due to partial volume effects and segmentation errors. $BV_j$ is thus computed analytically by integrating the inner lumen area $A_j(l)$ along length $L_j$ of the $j^{th}$ branch.

$$A_j(l) = \pi \left( \frac{d_j(l)}{2} \right)^2$$

$$= \frac{\pi}{4} (m_j \times l + c_j)^2$$

$$BV_j = \int_0^{L_j} A_j(l)\, dl$$

$$BV_j = \frac{\pi(L_j^3 {m_j}^2 + 3L_j^2 c_j m_j + 3L_j {c_j}^2)}{12} \quad \text{(Eq. 2)}$$

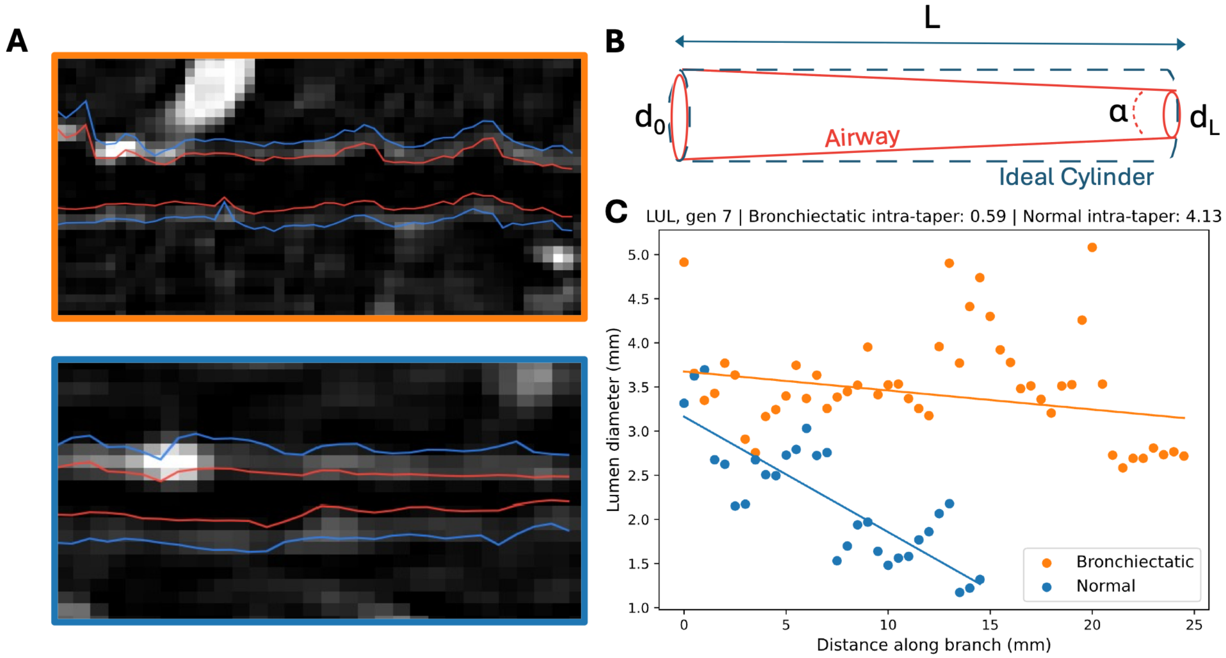

Figure S7: (A) Multiplanar reconstructed generation-7 airway branches from left upper lobe from the same patient showing a bronchiectatic airway (orange outline) and a tapered airway (blue outline). (B) Graphic explaining the concept of cylindricity (ratio of airway volume to an ideal cylinder of diameter $d_0$ and length L) and tapering angle α. (C) Lumen diameter along the length of the normal (blue) and bronchiectatic (orange) airway branch with Huber robust regression estimated regression lines.

**Luminal Surface Area ($SA_j$):** Assuming the airway branch as an open cylinder, surface area is computed by integrating the circumference of airway lumen along the length $L_j$of the $j^{th}$ branch. Thus, $SA_j$ is computed by integrating lumen circumference $\pi d_j(l) = \pi(m_j \times l + c_j)$ over $[0, L_j]$:

$$SA_j = \pi \int_0^{L_j} (m_j l + c_j)\, dl = \frac{\pi}{2}\left(L_j^2 m_j + 2L_j c_j\right) \quad \text{(Eq. 3)}$$

**Surface Area to Volume Ratio (SA:V ratio):** SA:V ratio was computed as the ratio of branch surface area computed using Eq. 3 to branch volume computed using Eq. 2

**Average Diameter ($\bar{d}_j$)** of a branch is analytically estimated as

$$\bar{d}_j = \frac{1}{L_j}\int_0^{L_j} d_j(l)\, dl = \frac{1}{L_j}\int_0^{L_j} (m_j l + c_j)\, dl = \frac{L_j m_j + 2c_j}{2} \quad \text{(Eq. 4)}$$

**Diameter Standard Deviation ($\sigma_j$):** Standard deviation of diameter along the branch is derived analytically as:

$$\mathrm{Var}[d_j] = \frac{1}{L_j}\int_0^{L_j} \left(m_j l + c_j - \bar{d}_j\right)^2 dl = \frac{{m_j}^2 {L_j}^2}{12}$$

$$\sigma_j = \sqrt{\mathrm{Var}[d_j]} = \frac{|m_j L_j|}{\sqrt{12}} \quad \text{(Eq. 5)}$$

**Intra-branch Taper ($\delta_j$):** is defined as the normalized gradient of airway diameter along an airway branch (Figure S7C):

$$\delta_j = -\frac{m_j}{c_j} \quad \text{(Eq. 6)}$$

Higher values of $\delta_j$ indicate progressive narrowing (normal tapering); lower values indicate dilation (bronchiectasis).

**Cylindricity ($C_j$)** of the $j^{th}$ airway branch is defined as the ratio of airway branch volume ($BV_j$) to the volume of an ideal cylinder ($CV_j$) of length $L_j$ and radius $c_0$ (Figure S7B), and is computed as:

$$C_j = \frac{BV_j}{CV_j} = \frac{BV_j}{\pi\left(c_j/2\right)^2 L_j} \quad \text{(Eq. 7)}$$

Ideal cylinder volume can be computed simply as $CV_j = \pi\left(c_j/2\right)^2 L_j$. Normal airways with progressive narrowing yield $C_j < 1$. Bronchiectatic airways approach $C_j = 1$ (cylindrical).

**Tapering Angle ($\alpha_j$)** reflects the degree of narrowing along the airway's length (Figure S7B) and is computed using the following equation

$$\alpha_j = \arctan\left(\frac{\left|c_j - d_j(L_j)\right|}{L_j}\right) = \arctan\left(\frac{\left|m_j L_j\right|}{L_j}\right) = \arctan\left|m_j\right| \quad \text{(Eq. 8)}$$

**Residual Zero-Crossing Rate ($ZCR_j$):** Let $r_j(l)$ be the regression residual obtained from Equation 1 at point $l$. Let branch $j$ of length $L_j$ consists of M points. The number of zero-crossings are:

$$ZC_j = \sum_{l=0}^{M} 1\left[r_j(l)\, r_j(l+1) < 0\right] \quad \text{(Eq. 9)}$$

$$ZCR_j = ZC_j/M \quad \text{(Eq. 10)}$$

**Inter-Branch Tapering ($\lambda_j$)** is defined as the relative change in median airway diameter with increase in airway generation

$$\lambda_j = \frac{\bar{d}_{\text{j-1}} - \bar{d}_j}{\bar{d}_{\text{j-1}}} \quad \text{(Eq. 11)}$$

Where $\bar{d}_{\text{j}}$ and $\bar{d}_{\text{j-1}}$ are the average diameters of the $j^{th}$ and its parent ($j - 1^{th}$) airway branch computed using Eq. 5, respectively.

The tapering in normal airways yields a cylindricity value less than 1 and a higher intra/inter- branch tapering, indicating progressive narrowing towards the distal end. Bronchiectasis disrupts this pattern, leading to cylindrical airways with cylindricity values approaching 1, and reduction in tapering indices. This change reflects a loss of normal tapering and a tendency towards uniform dilation. In suppurative diseases, bronchiectasis is often preceded by mucus deposition along an airway branch, which may occur in discontinuous segments rather than a continuous layer. This can result in increased diameter variability and frequent residual zero-crossings, signaling early disease progression before permanent structural changes sets in.

**S2.9.3: Architectural Distortion Features**

**Tortuosity** ($\tau_j$) is the ratio of arc length to the Euclidean chord length:

$$\tau_j = \frac{L_j}{\left\|p_j^{end} - p_j^{start}\right\|_2} \quad \text{(Eq. 12)}$$

Where $p_j^{start}, p_j^{end}$ are the starting and ending co-ordinates of the $j^{th}$ airway branch. Values of $\tau_j > 1$ indicate a tortuous branch; $\tau_j = 1$ is a straight branch.

**Curvature and Torsion:** Pointwise curvature and torsion were computed using the genepy3d[11] from the branch coordinates in physical space. Torsion was smoothed with a 1D Gaussian filter (σ = 1 sample). First-order statistics (mean, median, SD, IQR, min, max, skewness, kurtosis) were computed for each quantity across all M sampling points of a branch.

**Direction Angle:**

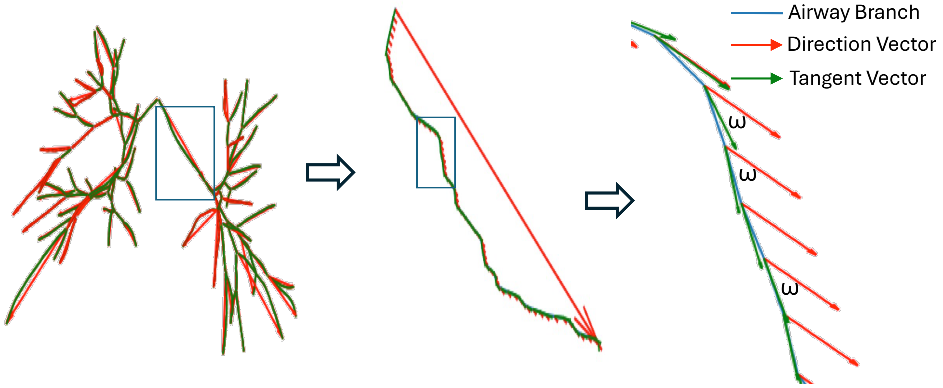

Figure S8: Direction angle (ω) measurements along each point on an airway branch (blue) computed as the angle between local tangent vector (green) and direction vector (red).

The direction vector of branch $j$ is:

$$d_j = \frac{p_j^{end} - p_j^{start}}{\left\| p_j^{end} - p_j^{start} \right\|_2}$$

At each sampling point $m$ along the branch, the direction angle[12] ($\omega_m$) is the angle between $d_j$ and the local tangent vector $t_m$ (Figure S8):

$$\omega_m = arccos\left( \frac{d_j \cdot t_m}{\left\| d_j \right\|_2 \left\| t_m \right\|_2} \right) \quad \text{(Eq. 13)}$$

First-order statistics of $\omega_k$ were computed per branch.

**Branching Angles:** The bifurcation angle between sibling branches $j$ and $j'$ sharing a common parent is:

$$\theta_{sib} = arccos\left( \frac{v_j \cdot v_j'}{\left\| v_j \right\|_2 \left\| v_j' \right\|_2} \right), \quad v_j = p_j^{end} - p_j^{start}$$

The parent-child angle $\theta_{par}$ is computed analogously using the vectors of branch $j$ and its parent branch $(j - 1)$.

**S2.9.4: Global Airway Descriptors**

**Graph-Theoretic Features:** Nine features were extracted from the undirected version of G (edge-weighted by branch arc length) for each region:

Table S1: Graph-Theoretic Airway Descriptors

| Feature | Definition |
|---|---|
| Graph diameter | Maximum shortest-path hop count between any two nodes |
| Average shortest path length | Mean Dijkstra path length (mm) over all node pairs |
| Graph density | Ratio of existing edges to the total maximum possible edges |
| Average clustering coefficient | Mean of local clustering coefficients |
| Transitivity | 3 × triangles / triads |
| Local efficiency | Mean of nodal efficiencies |
| Global efficiency | Mean of all pairwise inverse distances |
| Max adjacency eigenvalue | Spectral radius of the adjacency matrix |
| Wiener index | Sum of all pairwise shortest path lengths |

**Fractal Dimension:** The box-counting fractal dimension of the airway mask and skeleton was estimated separately using a log-spaced set of $N_s = 20$ box sizes $\varepsilon \in [\varepsilon_{min}, \varepsilon_{max}]$. Physical voxel coordinates were used to account for anisotropic spacing.

**Shape Features:** The airway mask for each region was cropped to its bounding box, and the following shape features were extracted using PyRadiomics[13]: mesh-based volume, voxel volume, surface area, surface-to-volume ratio, and maximum 3D and 2D diameters (row, column, slice directions).

**Carina-to-Endpoint Path Lengths:** For each lobe, the Dijkstra shortest path length (weighted by branch arc length) from the carina node $c^*$ to each terminal leaf node within that lobe was computed. Summary statistics (mean, median, SD, IQR, min, max, skewness, kurtosis) and imbalance (max − min, a measure of within-lobe tree asymmetry) were extracted.

#### S2.9.5: Feature Aggregation

Per-branch features (excluding branches with less than 4 valid cross-sectional measurements) were aggregated to the patient level by computing eight summary statistics – mean, median, SD, IQR, minimum, maximum, skewness, and kurtosis. This produced a 462-dimensional feature vector per patient for each of seven regions: overall tree (trachea, and main bronchi), LUL, LML, LLL, RUL, RML, and RLL. Lobar features were extracted if at least 3 airway branches were detected.

#### S3: Test-Retest Stability Analysis:

To assess the test-retest repeatability of quantitative airway measurements, the publicly available RIDER Test-Retest dataset with N=31 non-small cell lung cancer patients was used from The Cancer Imaging Archive (TCIA)[14,15]. These patients underwent same day repeat CT scans acquired within 15 minutes of each other under identical imaging conditions, providing paired scans suitable for evaluating measurement stability in the absence of true physiological change. This dataset was used exclusively for feature stability analysis and was not used for model training or evaluation.

To identify reproducible RadAr features suitable for downstream modeling, concordance correlation coefficient (CCC) was computed between paired test and retest CT scans from the RIDER dataset across 462 features derived from the overall airway tree. Features with CCC greater than 0.74 were categorized as highly stable, those with CCC greater than 0.49 as moderately stable, and all remaining features as unstable.

Of the 462 RadAr features extracted from the RIDER test-retest cohort, 227 (49.1%) met the minimum stability threshold of CCC > 0.49 and were retained for downstream analyses, of which 148 (32.0%) were highly stable (CCC > 0.74) and 79 (17.1%) moderately stable (0.49 < CCC ≤ 0.74). CCC values ranged from -0.54 to 0.97 with a median of 0.48, reflecting substantial heterogeneity across feature categories.

Overall, global tree-level descriptors and volumetric shape features were near-universally stable (surface area CCC = 0.972, voxel volume CCC = 0.968), reflecting their insensitivity to local segmentation or skeletonization variability. Airway dimensions were also stable at the mean and median level across lumen area, outer area, wall area, diameter, perimeter, eccentricity, elongation, and wall-to-lumen ratio, though distributional extremes (min, max) and shape statistics (skewness, kurtosis) were frequently unstable.

Central tendencies of tapering features (mean, median) were stable but became unreliable at distributional extremes (min, max, skewness, kurtosis). Tapering angle was the most broadly stable individual feature with mean, median, standard deviation, and IQR all meeting the threshold (CCC = 0.859, 0.695, 0.592, 0.641), while intra-branch tapering was the least reproducible, with only its mean and median moderately stable. Cylindricity mean was stable (CCC = 0.797) though spread and extreme statistics were not. Zero crossing mean (CCC = 0.868) and zero crossing rate mean and median (CCC = 0.730, 0.750) were reliably stable, indicating that residual-based diameter irregularity measures are reproducible despite their sensitivity to segmentation surface noise.

Architectural distortion features of curvature, torsion, and directional angle were the least reproducible features. Across all categories, mean and median statistics were consistently the most stable summary measures, while skewness, kurtosis, and extremes were systematically excluded. Tortuosity mean (CCC = 0.731) was stable but spread and extreme statistics were not. Curvature mean-level statistics showed borderline reproducibility across sub-families (CCC ≈ 0.73-0.76), while higher-order statistics were largely unreliable. Branching angle features were moderately stable at the mean level (parent angle CCC = 0.782, sibling angle CCC = 0.624) but not beyond. Torsion and directional angle features were effectively non-reproducible, with torsion achieving stability in only 3 of 64 cases, reflecting the high sensitivity of pointwise geometric derivatives to skeletal resampling and segmentation surface irregularities.

**S4: Classification Model Training and Evaluation**

Feature selection and model training were conducted within a Monte Carlo cross-validation (MCCV) framework with 300 repeats, which is analogous to bootstrap sampling without replacement and enables identification of low-variance, consistently selected features while providing stable estimates of model performance. First, features with pairwise Pearson correlation exceeding 0.95 were removed. From this reduced feature set, the top p features were selected using the Wilcoxon rank-sum test, subject to the constraint that the training set contained at least 10×p observations to avoid overfitting. Features consistently selected across the 300 cross-validation folds were used to train the final locked-down classification model. Z-score normalization parameters were derived exclusively from the training set and applied to cross-validation folds and all holdout or external test sets to prevent data leakage. For the COVID-19 cohorts, feature selection was performed on the subset of 227 stable features identified from the RIDER test-retest analysis. For the fibrotic ILD cohort, models were developed as part of the AIIB23 challenge prior to the stability analysis and LML labelling, and all 462 features were therefore used as input to the selection pipeline; the challenge submission portal has since closed, precluding re-evaluation of a stability-filtered and LML model.

Model performance was evaluated using balanced accuracy, sensitivity, specificity, and F1 score. For COVID-19, the area under the receiver operating characteristic curve (AUC) and average precision (AP) were also computed but were unavailable for the fILD dataset due to limitations of the challenge submission portal. To assess the independent value of RadAr for predicting COVID-19 severity, multivariable logistic regression models were fitted on $D_2^{ext}$ adjusting for systolic blood pressure, respiratory rate, heart rate, pulse oximetry below 90% (binary), and age. Odds ratios, 95% confidence intervals, and p-values were reported. Equivalent adjustment was not possible for STOIC COVID-19 ($D_2$) or the fILD ($D_1$) since clinical information was unavailable.

### S5: Multivariable analysis for COVID-19 severity prediction

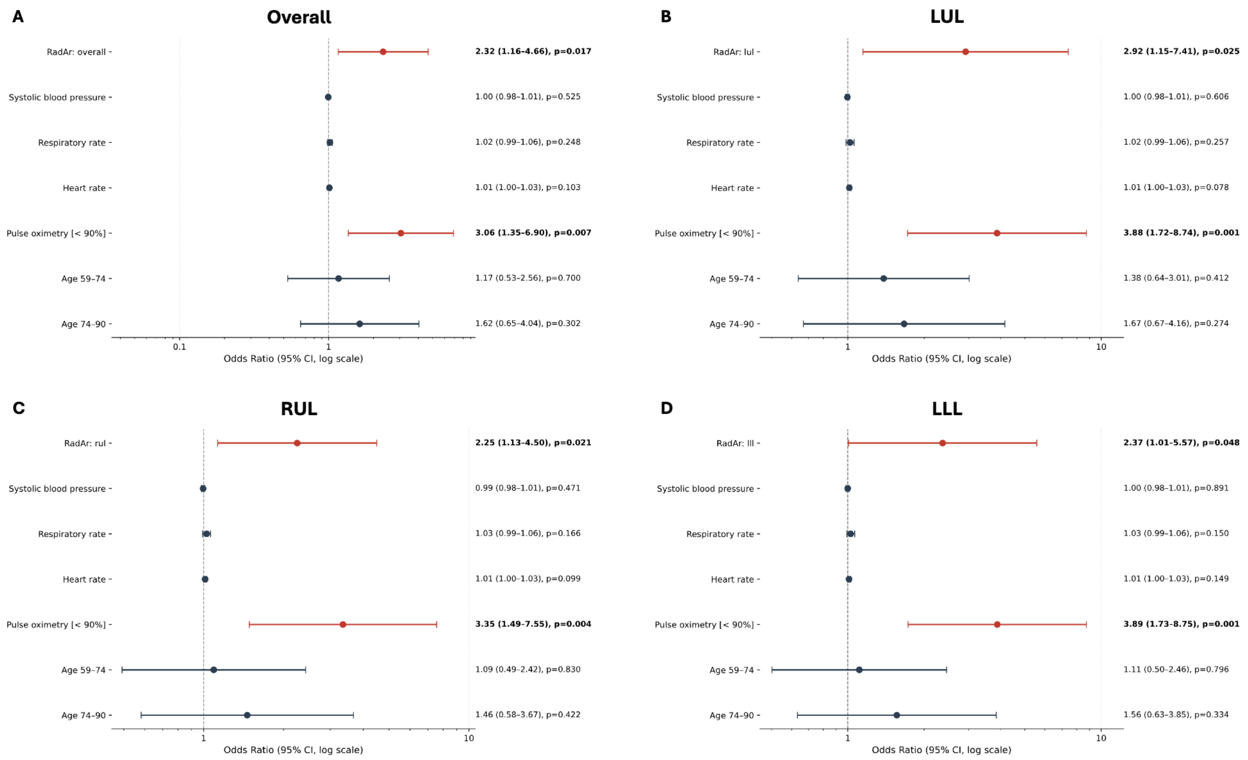

Figure S9: Multivariable logistic regression analyses for the COVID-19 severity prediction on $D_2^{ext}$ across (A) overall, (B) LUL, (C) RUL, and (D) LLL models.

### S6. Comparison of Median Branch SA:V Ratio with Overall Voxel Based SA:V Ratio in COVID-19

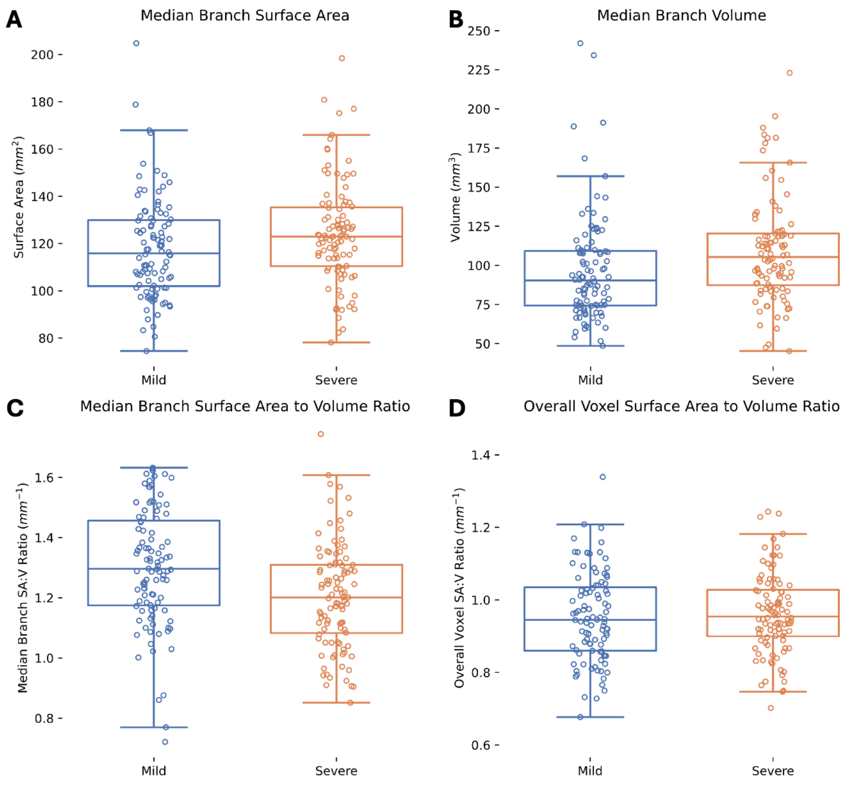

Figure S10: Box and strip plot for (A) median branch surface area, (B) median branch volume, (C) median branch surface area to volume ratio, and (D) surface area to volume ratio computed from overall airway mask through voxel based calculation in mild (blue) and severe (orange) COVID-19 patients from the training set ($D_2^{train}$).

### S7. Unsupervised Consensus Clustering

#### S7.1 Method

Of 227 repeatable features, highly correlated features (Pearson's |r|>0.90) were removed, retaining 116 of the features. Feature values were clipped to the corresponding within-dataset 1st and 99th percentiles, resulting in adjustment of 2.2% of all values, and were subsequently z-score standardized within each dataset. This procedure removed between-cohort differences in feature location and scale, thereby ensuring that clustering was driven primarily by relative morphological variation within each disease cohort rather than by acquisition related differences between datasets. Consequently, clustering reflects shared residual within-disease variation rather than disease separation.

We applied the Monti consensus clustering framework[16]. For each k=2-8, 250 subsampled were generated, each containing 75% of patients selected without replacement. Each sample was clustered using Ward agglomerative clustering. For every candidate k, the consensus matrix C recorded the proportion of subsamples in which each pair of patients was assigned to the same cluster. The optimal number of clusters was selected from the elbow of the area under the consensus cumulative distribution function curve and cross-checked using the proportion of ambiguous clustering, with optimal number of clusters identified at k=5. Final phenotype assignments were obtained by applying k-means clustering to the consensus matrix. The resulting phenotypes were labeled P1-P5 in ascending order of composite outcome rate (Table S2); outcome information was used only to order the phenotypes and did not contribute to their derivation.

Table S2: Distribution of clinical outcomes across consensus-derived airway phenotypes

| Phenotype | N | COVID-19 severe | fILD deceased |
|---|---|---|---|
| P1 | 461 | 20.2% (86/425) | 19.0% (4/21) |
| P2 | 338 | 22.2% (69/311) | 38.5% (10/26) |
| P3 | 315 | 28.6% (85/297) | 26.7% (4/15) |
| P4 | 201 | 33.1% (57/172) | 48.0% (12/25) |
| P5 | 156 | 31.1% (46/148) | 75.0% (6/8) |

**S7.2 Airway Morphology Across Consensus-Derived Phenotypes**

To characterize the structural basis of the five consensus-derived phenotypes, we compared seven representative overall-airway features across P1-P5. All seven features differed significantly across phenotypes (Figure S11). Since features were clipped and z-score standardized within each cohort, the standardized values provide the primary measure of between-phenotype contrast, whereas raw-unit medians are provided in for clinical interpretability (Table S3).

Feature analysis indicated the following airway morphologies across the five phenotypes:

**P1 (lowest risk):** Extensive, finely branched, thin-walled, non-dilated, and minimally tortuous airway tree.

**P2:** Compact, short, low-volume airway tree with relatively few branches and the greatest intra-branch tapering.

**P3:** Near-average global size with wall thickening and lumen narrowing, consistent with mild wall-predominant remodeling.

**P4:** Dilated, high-volume, heterogeneous branches with low SA:V and relatively thin walls; a large-airway, traction-bronchiectasis-like configuration.

**P5 (highest composite risk):** Sparse, dilated, thick-walled, and highly tortuous airway tree with low global extent but elevated minimum peripheral caliber and marked inter-branch variability.

Phenotypes P1 to P3 had relatively normal airways, whereas P4 and P5 showed structural alterations in the airway tree (Figure S12). Consequently, P4 and P5 had a larger number of composite events (severe COVID-19, mortality in fILD) as compared to P1-P3.

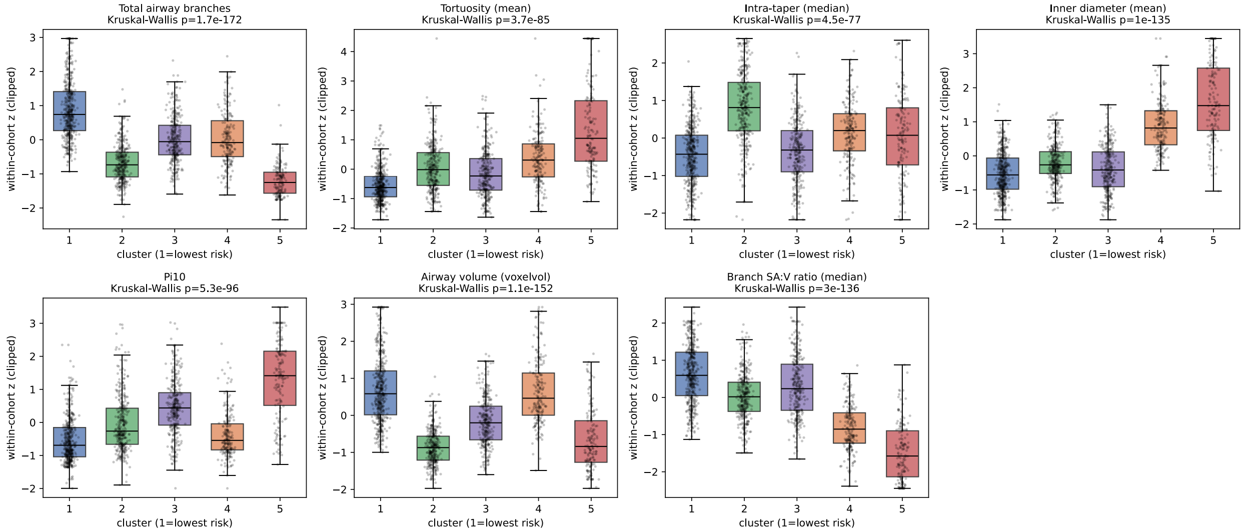

Figure S11: Distribution of within-cohort z-scored feature distribution across the five airway phenotype clusters

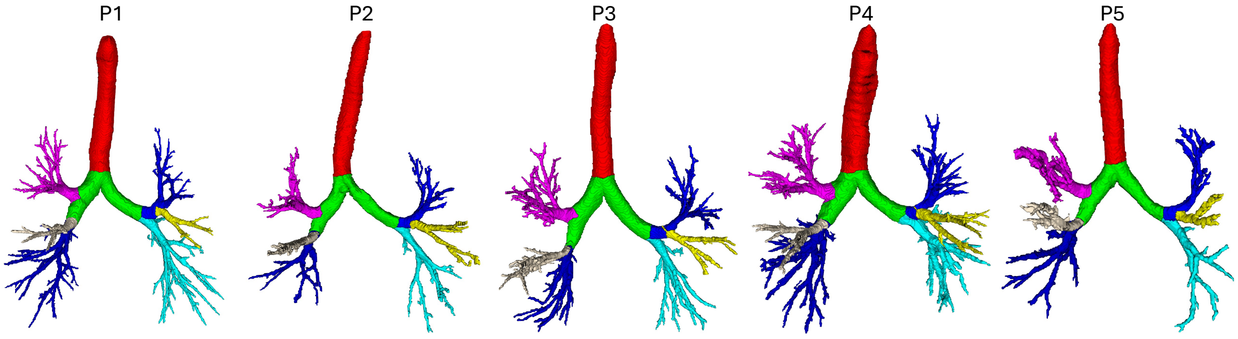

Figure S12: Representative examples of lobe-labelled airways across the five phenotypes

Table S3: Raw and within-cohort z-score standardized values of average overall-airway features across consensus-derived phenotypes.

| Feature | Unit | P1 | P2 | P3 | P4 | P5 | Kruskal-Wallis p |
|---|---|---|---|---|---|---|---|
| Total airway branches | z-score | **+0.88** | -0.68 | +0.01 | +0.03 | **-1.21** | p<0.0001 |
| | Raw | 217 | 104 | 155 | 154 | 67 | - |
| Tortuosity (mean) | z-score | -0.55 | +0.05 | -0.15 | +0.37 | **+1.35** | p<0.0001 |
| | Raw | 1.015 | 1.018 | 1.017 | 1.019 | 1.023 | - |
| Intra-taper (median) | z-score | -0.45 | **+0.84** | -0.35 | +0.14 | +0.05 | p<0.0001 |
| | Raw | 2.52 | 3.16 | 2.56 | 2.84 | 2.77 | - |
| Inner diameter (mean) | z-score | -0.52 | -0.20 | -0.39 | +0.89 | **+1.62** | p<0.0001 |
| | Raw (mm) | 3.33 | 3.53 | 3.42 | 4.20 | 4.54 | - |
| Pi10 | z-score | -0.55 | -0.04 | +0.45 | -0.39 | **+1.31** | p<0.0001 |
| | Raw (mm) | 4.62 | 5.01 | 5.79 | 4.72 | 6.98 | - |

| Feature | Unit | P1 | P2 | P3 | P4 | P5 | Kruskal-Wallis p |
|---|---|---|---|---|---|---|---|
| Airway volume | z-score | **+0.70** | -0.86 | -0.17 | +0.64 | -0.67 | p<0.0001 |
| | Raw ($mm^3$) | 40,064 | 20,697 | 29,508 | 39,350 | 21,036 | - |
| Branch SA:V ratio (median) | z-score | +0.63 | +0.03 | +0.30 | -0.81 | **-1.48** | p<0.0001 |
| | Raw ($mm^{-1}$) | 1.369 | 1.228 | 1.290 | 1.025 | 0.899 | - |

**S8: Pipeline Runtime**

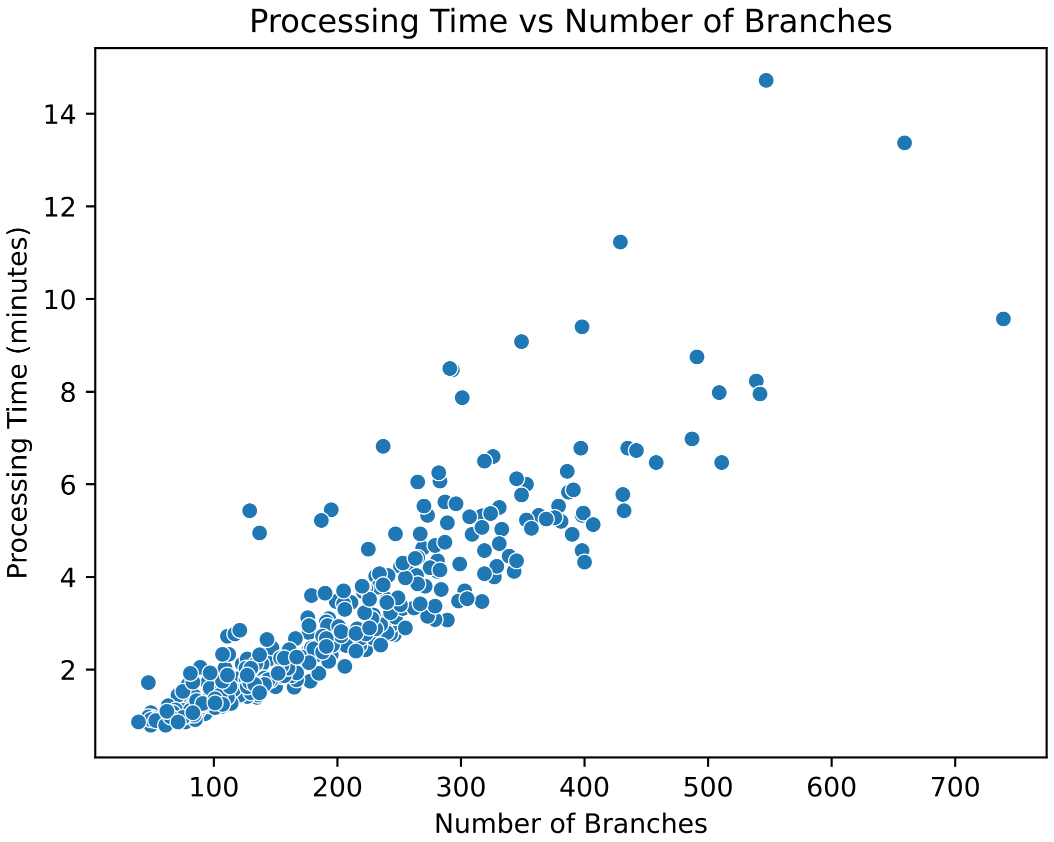

Figure S13: RadAr processing time in minutes as a function of number of segmented airway branches

Given the CT scan and associated airway segmentation, the computational efficiency of RadAr was assessed by recording end-to-end processing time per case across a subset of 358 patients. This included skeletonization, DAG generation, lobar labelling, cross-sectional plane interpolation, FWHM measurements, and feature extraction. The average runtime was 3.00 ± 2.03 minutes, with a median of 2.40 minutes and an interquartile range of 2.47 minutes. Majority of cases completed in under 4 minutes (minimum 0.8 minutes) and a small tail extending to a maximum of 14.72 minutes. Processing time scaled with the number of segmented airway branches, with cases at the upper extreme corresponding to large, densely segmented airway trees in which a greater number of cross-sections and per-branch features were computed (Figure S13). All timings were measured on a single CPU thread without GPU acceleration; the runtime can be further reduced through parallelization across cases, which is naturally supported by the pipeline architecture.
